# The Failures of an Ideal COVID-19 Vaccine: A Simulation Study

**DOI:** 10.1101/2021.11.22.21266669

**Authors:** Robert J. Kosinski

**Affiliations:** Emeritus of Biological Sciences, Clemson University

## Abstract

This paper simulates an ideal COVID-19 vaccine that confers immediate sterilizing immunity against all SARS-CoV-2 variants. The purpose was to explore how well this ideal vaccine could protect a population against common conditions (such as vaccine hesitancy) that might impair vaccine effectiveness. Simulations were done with an SEIRS spreadsheet model that ran two parallel subpopulations: one that accepted vaccination, and another that refused it. The two subpopulations could transmit infections to one another. Success was judged by the rate of new cases in the period from 1-5 years after the introduction of the vaccine.

Under good conditions, including a small subpopulation that refused vaccination, rapid distribution of the vaccine, duration of vaccinal immunity longer than 12 months, good retention of interest in getting vaccinated after the first year, strict maintenance of nonpharmaceutical interventions (NPIs) such as masking, and new variants with R_0_s less than 4.0, the vaccine was able to end the epidemic. With violation of these conditions, the post-vaccine era futures ranged from endemic COVID at a low or medium level to rates of COVID cases worse than anything seen in the US up to late 2021. The most important conditions for keeping case rates low were a fast speed of vaccine distribution, a low percentage of the population that refuses vaccination, a long duration of vaccinal immunity, and continuing maintenance of NPIs after vaccination began. On the other hand, a short duration of vaccinal immunity, abandonment of NPIs, and new variants with a high R_0_ were powerful barriers to disease control. New variants with high R_0_s were particularly damaging, producing high case rates except when vaccination speed was unrealistically rapid. A recurring finding was that most disease afflicting the vaccinated population in these simulations originated in the unvaccinated population, and cutting off interaction with the unvaccinated population caused a sharp drop in the case rate of the vaccinated population.

In conclusion, multiple common conditions can compromise the effectiveness of even an “ideal” vaccine.

## Introduction

The production of vaccines against the SARS-CoV-2 virus was a landmark development in a pandemic that has caused over 44 million cases and taken over 764,000 lives in the US as of 18 November 2021 (CDC, 2021a). Vaccines can be a powerful public health tool against COVID-19. Moutinho (2021) described how the town of Serrana, Brazil vaccinated 27,000 of the 45,000 residents with CoronaVac, and achieved an 80% drop in symptomatic COVID cases. Balicer and Ohana (2021) reported that Israel achieved a 100-fold drop in new cases by an energetic campaign that vaccinated 2.5% of the population on some days. However, then they warned that this rapid Israeli progress may be encountering obstacles like vaccine hesitancy and impatience with non-pharmaceutical interventions (NPIs) such as masking.

Similar vaccine problems are occurring in the US. As of 17 November 2021, 58.9% of the US population had been fully vaccinated (CDC, 2021a), and booster doses were being given to certain populations. The rate of completed vaccinations peaked at about 2.5 million/day on 8 April 2021 (reaching 0.76% of the whole population on that one day). However, by mid-November, the daily rate was less than 300,000 completed vaccinations (about 0.09%/day) (CDC, 2021b). The average rate was between these limits. If we assume that vaccinations in the US began on 14 December 2020, and that the target population was solely those of the age of 12 or older, the average rate of exponential decline in the unvaccinated fraction of this 12+ population by mid-November was 0.34%/day over 338 days. The average rate for the whole population over the same time period was 0.26%/day.

Although the quick development of high-efficacy vaccines was a scientific triumph (Golob, 2021), producing the vaccine is only part of the public health battle. Aschwanden (2021) summarized reasons why herd immunity would be difficult to achieve through vaccination: Vaccines might still allow disease transmission, vaccine rollouts in different countries have been slow and uneven, there was (at the time) still no vaccine for children, pockets of vaccine hesitancy would serve as reservoirs of disease, new variants will develop, vaccine-induced immunity will expire, and the arrival of vaccines might cause the public to discontinue NPIs such as masking. Murray and Pitot (2021) also raised these concerns, and added that winter COVID outbreaks may become the norm in the future.

Given all these limitations, what is the best COVID control that we could expect from an excellent vaccine?

### This Report

In this paper’s model scenario, an epidemic starts when an infected individual enters a population at 0 years, and the epidemic proceeds with control only by NPIs for a year. At that point, an “ideal” vaccine is developed and distributed. This vaccine:

a. produces same-day, sterilizing immunity against all variants;
b. renews the immunity of recipients that already have disease-induced immunity; and
c. induces vaccinal immunity that lasts from 6 to 24 months. During these months, no vaccinated individual can be reinfected.

The success of the vaccine is judged by the “post-vaccine case rate,” or the number of COVID cases per thousand per year over the period from one year to five years after the introduction of the vaccine. The more the case rate was reduced over these four post-vaccine years, the more effective the vaccination regime was judged to be. Anthony Fauci has suggested in interviews that the US should aspire to fewer than 10,000 infections/day (Schreiber, 2021). This equates to approximately 10 cases per thousand per year, giving an average individual a 1% chance of contracting COVID-19 each year. Therefore, an average post-vaccine case rate of 10 cases/thousand/year or lower will be regarded as a success.

In rough terms the model simulates the US experience of 2020 (initial outbreak controlled by NPIs only), introduction of the vaccine and the initial vaccination campaign in 2021, and then the four-year “post-vaccine era” of 2022-2025. It draws its conclusions from case rates in the “2022-2025” time period. Its emphasis is on the long-term, post-vaccine COVID environment.

Because the simulated vaccine is more effective than any real vaccine, it might be said that what this imaginary vaccine *can* accomplish is of little interest, but what it *cannot* accomplish is the real point. Plausible conditions that would make the “ideal” vaccine ineffective would probably make *any* vaccine ineffective.

The variables tested for their effect on the post-vaccine case rate were:

a. the speed of vaccine distribution, measured by the percent of susceptibles and disease-induced immunes vaccinated per day (0% to 1%);
b. the duration of vaccine-induced immunity (6, 9, 12, 18, or 24 months);
c. the percent of the population willing to accept vaccination (25% to 100%);
d. the year-to-year persistence of public interest in getting vaccinated;
e. abandoning or weakening NPIs such as masking once vaccinations become common;
f. the social isolation of the vaccinated and unvaccinated populations from each other (ranging from random mixing to total segregation); and
g. the development of a new variant with a higher R_0_ (changing from a “2020” base value of 2.3 to “2021” values of 3.0, 4.0, 5.0 or 6.0).

### Past COVID Vaccination Simulation Studies

Many COVID simulation studies have investigated the strategy of vaccination campaigns. For example, should the main target of the vaccination campaign be those that primarily spread the disease (young people), or those who suffer the most from the infection (older people)? If there is a two-vaccine treatment, how long should the interval between the two doses be? How can scheduling of doses minimize the development of vaccine-resistant variants? These are important questions, but are not the focus of this paper. This paper investigates the ability of vaccination to control an epidemic in the post-vaccine era.

Saad-Roy *et al*. (2020) investigated some of the same questions considered by this study. Using a model that allowed reinfection after immunity had expired, they simulated the five-year course of COVID in a northern, temperate location such as New York, with seasonal peaks in the fall, partial suppression of spread by non-pharmaceutical interventions (NPIs), and the inception of vaccination 1.5 years after the beginning of the epidemic. They used vaccination rates less than 1% per week and durations of vaccine-induced immunity of between 0.25 and 2 years. They found that high vaccine efficacy, fast rates of vaccine administration, and long durations of vaccinal immunity had the greatest chance of ending the epidemic. They emphasized the importance of vaccination speed, not just coverage and efficacy. They found that vaccine “refusers” could reduce overall immunity and allow outbreaks despite vaccination.

Paltiel *et al*. (2020) modeled vaccination campaigns with an SEIR model, using vaccines that were disease-modifying, disease-preventing, and a combination of these. Paltiel *et al*. emphasized “pace” of vaccinations in addition to efficacy and coverage. They concluded that a rapid rate of vaccine administration might be more important than the efficacy of the vaccine. Their base pace was vaccination of 0.5% of the population per day, but values of up to 2% per day were tested. They also assumed a linear completion of vaccination, so that to vaccinate 50% of the population would take only 100 days at a constant 0.5%/day. They warned that maintenance of NPIs such as masking would be vital even after the vaccine was introduced. Because they only simulated for six months, they assumed that expiration of immunity would not be a factor.

Patel *et al*. (2021) described an agent-based model of a vaccination campaign in North Carolina, including different races and ethnicities, and simulated 75% vaccine coverage of the population in six months, although they conceded that this vaccination rate was optimistic. They found that high vaccine coverage with a low-efficacy vaccine was better than low coverage with a high-efficacy vaccine. They did not include expiration of immunity in their simulations, and they found that abandoning NPIs while the vaccines are distributed “may result in substantial increases in infections, hospitalizations, and deaths.”

Gumel *et al*. (2021) simulated an SIR model in which there was no exit from the “removed” compartment (meaning that immunity did not expire), but in which new susceptibles were constantly being supplied by births. They anticipated that vaccination might end the epidemic, but that then COVID-19 would enter an endemic phase. They concluded that elimination of COVID-19 from the US was a practical goal, but warned that NPIs would have to be used in addition. An interesting feature of their model was that they divided the population into those who did and did not wear face masks. Disease control was most likely when the face mask group was larger and some of the other group started to wear masks. They predicted that herd immunity would be able to end the pandemic in the US in 2021, but only if NPIs are increased and sustained.

Moghadas *et al*. (2021) described an agent-based model of the US that predicted a vaccine with a maximum 70% coverage in any age group (delivered at about 2% of the population per week) could substantially reduce hospitalizations and death, even if the vaccine had only limited ability to prevent infection. They cautioned that maintenance of NPIs would be necessary to achieve these effects. However, they assumed that immunity would not expire before the end of their simulation (300 days).

Olivera Mesa *et al*. (2021) simulated the effect of vaccine hesitancy and relaxation of NPIs in the US, France, Germany, and the UK. They found that with a high-efficacy vaccine and a fast rate of vaccine distribution (10 months to vaccinate the whole population over 15 years of age), NPIs could be lifted when the vaccination campaign was finished. However, with some of the population refusing to get vaccinated and a vaccine with a lower efficacy, termination of NPIs led to periodic outbreaks whose period was determined by the duration of immunity. They concluded that reducing vaccine hesitancy had to be a primary public health goal.

Sandmann *et al*. (2021) evaluated the costs and successes of hypothetical vaccination programs in the United Kingdom, and included expiration of immunity in their model. They found that vaccination programs that produced a long duration of immunity (3 years) were much more successful than those with a shorter duration of immunity (45 weeks). They concluded that vaccination was well worth the cost, but prolonged use of NPIs was not.

Ke *et al*. (2021) estimated a COVID R_0_ of 5.8 for the US. Consequently, they estimated that 85-90% of the US population would have to be vaccinated to achieve herd immunity. They warned that efficacious and widely-distributed vaccines would be necessary for control, and that immunity against SARS-CoV-2 probably was not long-lived. They said that if immunity lasts on the average 45 weeks to one year and the distributions of immunity durations are Gaussian rather than uniform, vaccinations will have to occur every few months.

Li and Giabenelli (2021) used an agent-based COVASIM model to simulate the first six months of the COVID epidemic. They pointed out that vaccines might have the counterintuitive effect of temporarily raising case rates. This would occur because if old people are vaccinated first, vaccines will have little effect on reducing the spread of the disease (mainly done by the young), but might cause some to abandon NPIs. They concluded that vaccines alone cannot end the pandemic; NPIs must be continued as well.

Sah *et al*. (2021) simulated using an agent-based model of the US, and emphasized the importance of speedy distribution of the vaccine, mainly to forestall the appearance of more transmissible variants. The more transmissible the new variants were, the more important fast vaccine distribution became.

Betti *et al*. (2021) simulated COVID with and without vaccination out to 2022 in Ontario, and found that vaccination could substantially reduce disease incidence, but that if NPIs were abandoned too early, the benefits of vaccination would be lost. Their model did not include expiration of immunity before the simulation was finished.

Alagoz *et al*., 2021 used an agent-based model to predict COVID cases at two locations in Wisconsin and in New York City from February to December, 2021. Because of the short time period simulated, they assumed that vaccinal immunity would not expire. They used vaccination rates and NPI adherence to predict the “controllable spread date,” or the date at which new cases dropped below a certain fraction of the population. They found that vaccines were more effective when NPI adherence was high, and that a lack of adherence to NPIs would require vaccines to have a greater coverage and efficacy.

Mancuso *et al*. (2021) simulated the success of vaccines against new COVID-19 variants in the US out to 2024. Like this study, they divided the population into those who would and would not accept the vaccine. However, their model did not allow individuals who achieved immunity to be reinfected. They found that the more transmissible the new variant was and the lower the efficacy of the vaccine against it, the higher the portion of the population that needed to be vaccinated to achieve herd immunity. With the Pfizer and Moderna vaccines, they predicted a herd immunity threshold from 59% with the SARS-CoV-2 “wild-type” strain to 82% with the delta strain.

Truszkowska *et al*. (2021) simulated the city of New Rochelle, NY in great detail. They explored the tension between vaccination and reopening, both expressed as percent per day. The major conclusion was that reopening too fast would result in increases in infections and deaths. They recommended a vaccination rate of 1%/day.

Gozzi *et al*. (2021) simulated different vaccination strategies in six diverse countries ranging from Egypt to Canada. Vaccinal immunity did not expire during the length of their simulations. Their focus was on the role of NPIs as vaccinations were rolled out. They concluded that the “great effort” of vaccination would be wasted if NPIs were abandoned too soon. As a matter of fact, a slow vaccination speed could have the paradoxical effect of increasing deaths because it was ineffectual in conferring immunity, but still caused the public to abandon NPIs.

Several themes emerge from the papers above. To control COVID-19 with vaccination, the speed of vaccine distribution should be fast, vaccine efficacy should be high, and vaccine hesitancy should be low. NPIs should be maintained throughout the vaccine era. Finally, transmissible new variants pose a serious threat. This paper will echo all these themes.

## Methods

Note: Some of the model description below closely follows the same model description in Kosinski (2020).

The following paragraphs describe the model without vaccination or NPIs first, then the model’s simulation of NPIs, and finally the simulation of vaccination.

### The Simulation of an Unmitigated Epidemic

The simulations used an SEIRS model done using difference equations on a Microsoft Excel spreadsheet. The iteration interval (each line and column on the spreadsheet) was one day. The SEIRS model contained five different groups of individuals in a model population of one million: susceptible, exposed (infected but not yet infective), infective, disease-induced immunes (Diim), and vaccine-induced immunes (Viim). The overall model structure was a cycling of individuals between susceptibles and the two kinds of immunes:

The durations of these stages were as follows:

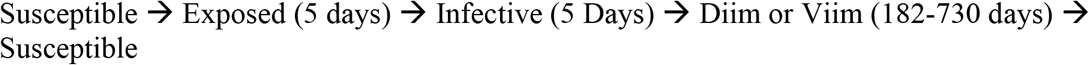

Both the Diim and Viim populations had the same duration of immunity. In an unmitigated epidemic or one mitigated only by NPIs, there would be no Viim individuals.

Each exposed, infective, and immune individual moved one day at a time through its compartment. For example, an exposed individual was moved from day one of exposure to day two, then to day three, etc. Although “Exposed” and “Infective” groups were handled separately in the calculations, they were sometime lumped into one group called “Infected” in the graphs that follow.

If we consider only one population and ignore corrections for use of NPIs (Eqs. 9 and 10), the number of new cases per day was given by

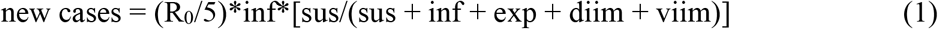

where inf = the number of infectives, exp is the number of individuals who are exposed to the disease but not yet infective, sus = the number of susceptibles, diim = disease-induced immunes, and viim = vaccine-induced immunes. The “5” refers to the five-day infective period. R_0_ was assumed to be 2.3 in the first set of simulations.

If we define the proportion of susceptibles in Eq. 1 (the quantity in brackets) as “psus” and rewrite the equation so that it refers to the number of new cases *per infective* for the whole five-day infective period, Eq. 1 becomes

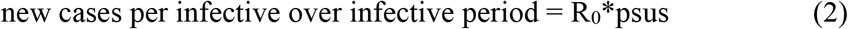

Therefore, when the fraction of susceptibles in a population is less than 1/R_0_, the new cases per infective are less than 1.0 during the whole infective period, and each infective will not replace itself during its five-day lifespan. Consequently, new cases per infective must decline. This becomes vital when we simulate the attainment of herd immunity. Herd immunity exists where each infective produces one or fewer new cases during its infective career, the epidemic cannot grow, and susceptibles can be largely protected from new infection. Persistent herd immunity is the objective of most vaccination campaigns.

The model had the capability of running two populations (accepting and refusing vaccination) on two different sheets. These were then combined on a third sheet. This allowed simulation of two populations that had different characteristics, but which interacted with each other by “sharing” infective individuals. The two populations could interact fully or partially. For example, only partial interaction might occur if the two populations lived in different areas or had reduced social interaction with one another. The degree of interaction was defined by an “interac” variable that was 1.0 if population 1 interacted just as much *per capita* with population 2 as with other members of population 1. If interac was less than 1, population 1 interacted less *per capita* with population 2 than with other members of population 1. If interac was zero, population 2 did not have any influence on population 1. There were analogous equations for population 2, but the interac variable was the same for both populations.

Let the total number of individuals in populations 1 and 2 be pop_1_ and pop_2_. Where the number of infectives in population 1 in an iteration is inf_1_ and the number of infectives in population 2 is inf_2_, then the number of infectives “seen” by populations 1 and 2 is

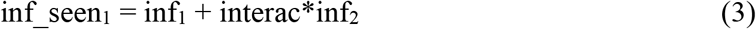

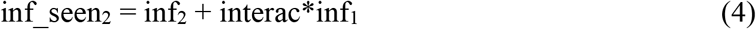

The importance of the proportion of susceptibles (psus) was seen in Eq. 2. The proportion of susceptibles in Population 1 and Population 2 will be

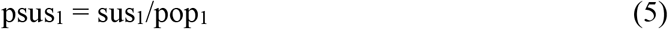

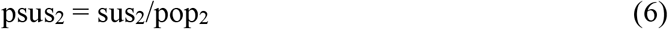

Two other variables, not to be confused with psus_1_ and psus_2_, are susfrac_1_ and susfrac_2_. These are the fraction of all the susceptibles that are in Population 1 and in Population 2. For example, say that at the beginning of the simulation, both Population 1 and Population 2 are all susceptibles, but ¾ of all the individuals are in Population 1. Then both psus_1_ and psus_2_ = 1.0, but susfrac_1_ = 0.75 and susfrac_2_ = 0.25. In other words,

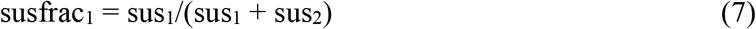

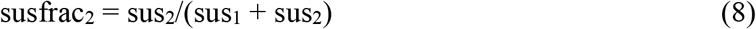

Ignoring the factor for social distancing for now, the number of new infections in a day on in populations 1 and 2 will be

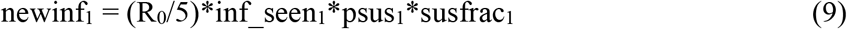

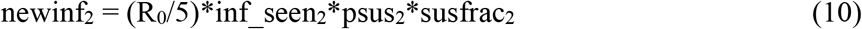

where the “5” refers to the five-day infective period. The “psus” factor reduces the number of new cases as the proportion of susceptibles in populations 1 and 2 falls, and will eventually reach zero if there are no susceptibles left in a population. But early in the epidemic, psus_1_ and psus_2_ will be close to 1.0, and every infective in populations 1 and 2 will produce R_0_/5 new cases per day. The susfrac variable allocates the new infections to Population 1 and Population 2, depending on what fraction of all the susceptibles each population had.

New infections delete members from the susceptible compartment and add them to day 1 of the exposed compartment. They then proceed through the exposed, infective, and disease-induced immune compartments, day by day, until they return to the susceptibles after the period of immunity has expired. Vaccination takes a fraction of the susceptibles and disease-induced immune compartments each day and deposits them in day 1 of the vaccine-induced immunity compartment. These individuals proceed through the number of days of vaccine-induced immunity and then return to the susceptible compartment. Vaccination is explained more fully below.

There were no death, birth, or demographic groups in the model (aside from Population 1 and Population 2). There was no seasonal change in infectivity. A single-population simulation started when one individual at the very beginning of its five-day infective career entered the population of 999,999 susceptible individuals. If two interacting populations were being used, each population got one infective on the first day. The sizes of Populations 1 and 2 were adjusted so their sum was 1,000,000 (e.g., if Population 1 was 750,000, Population 2 was 250,000).

### Exponential vs. Age-Structured Immunity

Probably the most distinctive aspect of the model was its treatment of the immune compartment. The traditional SEIRS model treats all the immune individuals as one pool from which a certain fraction (∂) leaves each iteration to re-join the susceptibles. If there were no new inputs into the immune compartment, this means that the immune population would experience an exponential decline, and that the average duration of immunity would be 1/∂. For example, for one year of immunity, 1/365 of the immunes would leave the immune compartment each day, and the average duration of immunity would be 365 days. I call this the “exponential” model of immune decline. However, since an exponential decline never reaches zero, some individuals can persist in the immune compartment indefinitely. For example, if we establish a cohort of 100 newly-immune organisms with a nominal length of immunity of one year, at one year the compartment will still contain about 37% of the initial cohort, about 14% after two years, and about 5% after three years. The exponential model will be used in a few cases in this report.

However, most of the results below use what might be called the “age-structured” model of immune decline. If the duration of immunity is set at one year, an individual enters the immune compartment and is moved one day at a time through 365 days of immunity. All the individuals who became immune 365 days ago will exit the immune compartment on the same day. Age-structured immunity allows the immune compartment to empty completely, and tends to cause fluctuations rather than equilibrium in the size of the immune compartment.

The duration of immunity had a major influence on the number of infections *per capita*. Durations of 6, 9, 12, 18, and 24 months of immunity were tested.

### Non-pharmaceutical Interventions

In the first year of each simulation, there was no vaccination, and all control of the epidemic relied on strategies like social distancing, masking, and hand-washing (NPIs). NPIs were simulated by multiplying R_0_ by (1 - lkd). Lkd was a “lockdown” factor that reached a maximum of 0.5. R_0_ was nearly undiminished when the infected percent of the population was low and reached 0.5*R_0_ when the infection rate was high. Lkd followed Michaelis-Menten kinetics (Eqs. 11 and 12). For population 1,

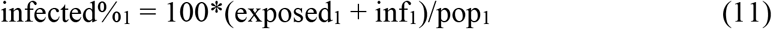

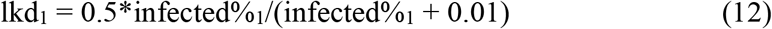

There were similar equations for population 2. In most simulations, populations 1 and 2 had the same lockdown equations and the same maximum lockdown parameter of 0.5.

Eq. 12 allowed the disease to spread rapidly at first, but then be controlled at a moderate level once the infected percent of the population increased beyond 0.01%.

### Vaccination

Population 2 was never vaccinated. In Population 1, a percentage (ranging from 0%-1% per day) of the susceptibles and of those with disease-induced immunity were moved into day 1 of the vaccine-induced immune (viim) population every day. Vaccination rates used in the model were 0%, 0.1%, 0.25%, 0.5%, and 1.0%/day.

The vaccine created same-day, sterilizing immunity against all variants, and could reset the immunity of those who already had disease-induced immunity to day 1 of the duration of immunity.

A number of vaccine doses were provided at the beginning of each year (starting one year into the simulation), and then were used up as vaccinations proceeded. If the number of available doses dropped to zero, vaccination stopped until the next “delivery” of doses arrived. If any doses were left at the end of the year, they were added to the next year’s vaccine supply, so no doses were discarded. The whole population might not be vaccinated in a year either because the number of doses was insufficient or because the vaccination rate was too slow to vaccinate all of Population 1 in a year. Therefore, while the supply of vaccine lasted, vaccination produced an exponential decline in the number of individuals in the susceptible and diim compartments. Note that this is not a linear decline in which a vaccination rate of 0.5% per day would be able to vaccinate a whole population in 200 days. If 0.5% of Population 1 is vaccinated per day, 84% of Population 1 could be vaccinated in a year; if 1% were vaccinated per day, 97% could be vaccinated in a year.

The number of vaccine doses supplied at the beginning of each year could be the same in successive years, or the number could decrease by 25%, 50%, 75%, or 100% per year to simulate the fading of interest in getting another vaccine. However, in all cases, the full number of doses to cover Population 1 was supplied in the first vaccine year.

Because the advent of the vaccine could reduce the willingness of the population to maintain NPI use, at 500 days since the start of the simulation (about “15 May” of the second year), the “lockdown” parameter either remained at 0.5, was reduced to 0.25, or was reduced to zero (complete abandonment of NPIs). This was meant to simulate the CDC announcement on 13 May 2021 that vaccinated individuals no longer needed to wear masks indoors or social-distance from others, which many feared would encourage the unvaccinated to do likewise.

All of the simulations above were done first with a pathogen with a constant R_0_ of 2.3 because inclusion of a new and more transmissible variant had such a powerful influence that it tended to obscure every other effect. After the “2.3” simulations, a reduced set of simulations was done with pathogens that started with an R_0_ of 2.3, but then increased their R_0_ to 3.0, 4.0, 5.0, or 6.0 between 500 to 600 days (“15 May” to “23 August” of the first year after vaccine introduction). For example, if the new R_0_ was to be 4.0, starting at 500 days, the quantity 0.01*(4.0 - 2.3) was added to the R_0_ until it reached 4.0 at 600 days, and then it was held at 4.0 for the rest of the simulation.

## Results

### General Simulation Outcomes

Before getting to the case rate results, we should contrast the typical course of an unmitigated epidemic, the effect of NPIs on this outcome, and then the additional effect of vaccination.

The three curves in Fig. 2 plot the percent of infected individuals (both exposed and infective) in three different epidemics, all with a constant R_0_ of 2.3 and a duration of immunity of one year. The blue curve represents a completely unmitigated epidemic. Infection there rose to maximum of 27% of the population at each peak, but any points above 2% are not shown in Fig. 2 in order to avoid compressing the “NPIs” and “Vac + NPIs” curves. The green curve and the orange curve employed NPIs that could produce a 50% reduction in R_0_ when the percent of the population infected was high. These NPIs were active from the beginning of the simulation. In addition to the NPIs, the green curve also used vaccination (delivered at 0.5% of the population per day) starting one year after the first case. All members of the population accepted the vaccine. The oscillations in the blue and orange curves are driven by the periodic expiration of immunity.

**Fig. 1.**
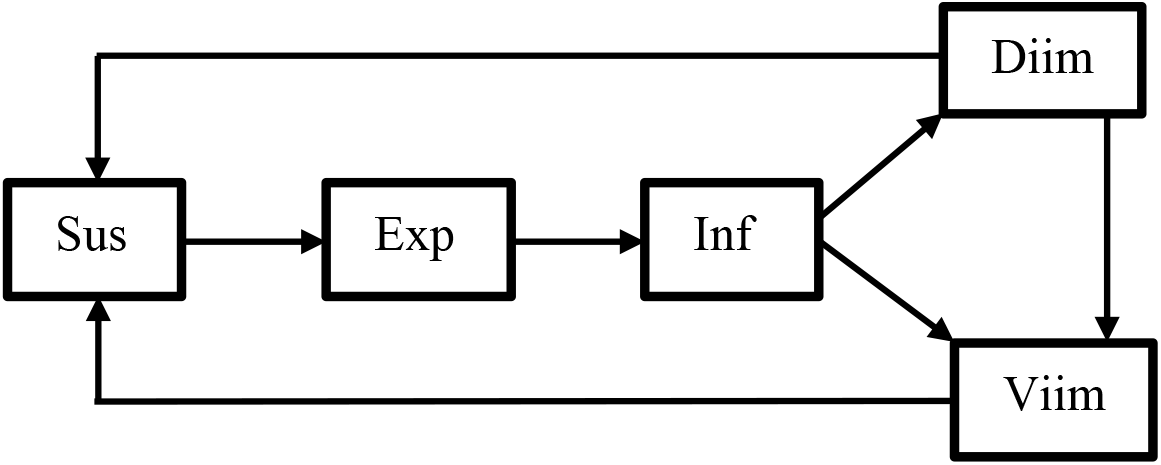
Flow of individuals in the model. The abbreviations stand for susceptibles (Sus), exposed (Exp), infectives (Inf), disease-induced immunes (Diim) and vaccination-induced immunes (Viim). Vaccination of disease-induced immunes could transfer them to the vaccine-induced immunes and reset their immunity to day one of the duration of immunity.

**Figure 2.**
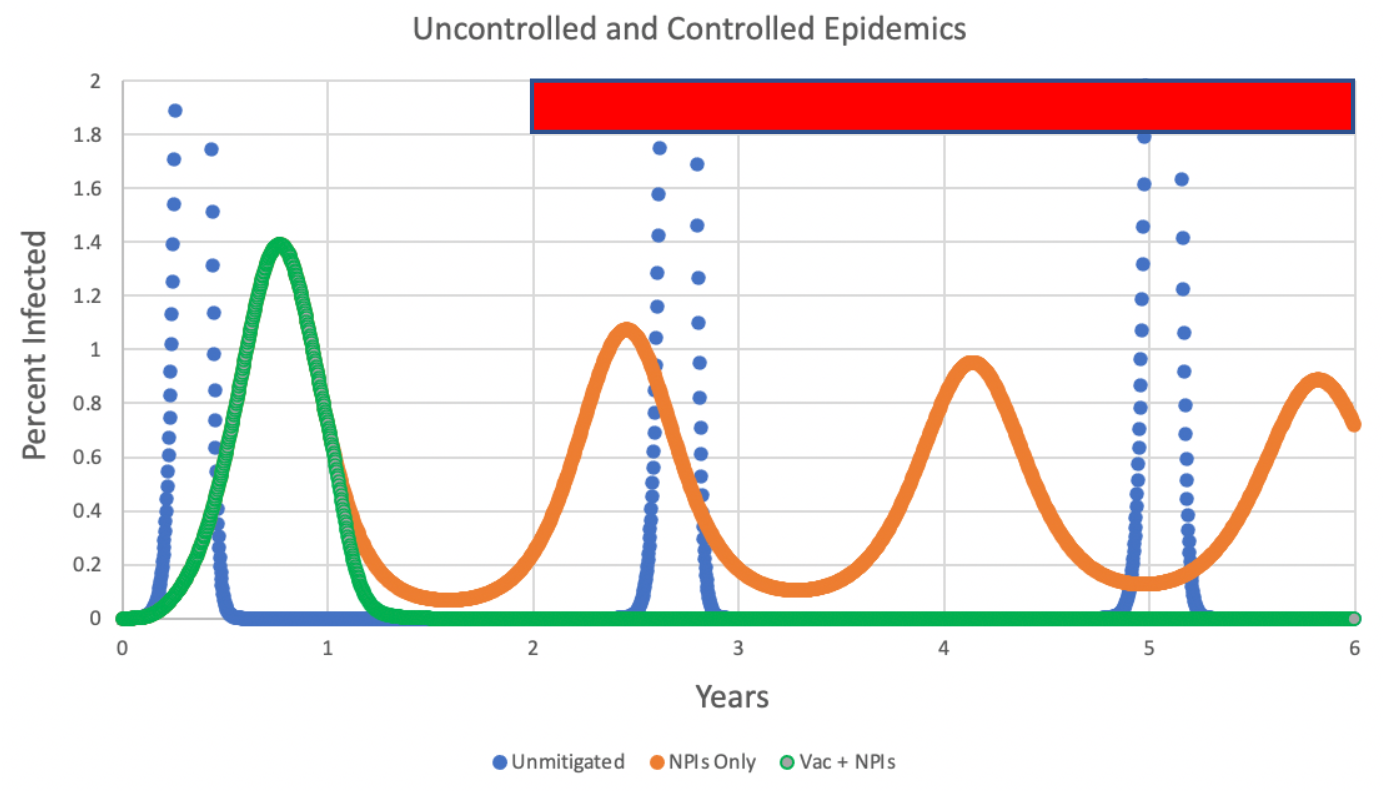
Typical results for R_0_ = 2.3 and one year of immunity for simulations of an unmitigated epidemic (blue), an epidemic controlled only by NPIs such as masking (orange), and an epidemic controlled by NPIs and also by vaccination that starts at one year (green). The red rectangle indicates the “post-vaccine era” in which case rates were computed.

The red rectangle in Figure 2 shows the “post-vaccine era” during which case rates were computed in order to compare the effect of vaccination regimes. This four-year period was used whether or not the simulations included vaccination.

The differences in the curves are striking. The unmitigated epidemic (blue curve) produced a seroprevalence at one year of 87% and a case rate during the time period encompassed by the red rectangle of 435.13 cases per thousand per year. Repeated peaks were caused by expiration of disease-induced immunity. Using NPIs alone (orange curve) reduced the one-year seroprevalence to 22% and the post-vaccine case rate to 159.83 cases per thousand/year over the last four years. Again, expiration of disease-induced immunity triggered much smaller oscillations than in the case of the unmitigated epidemic. However, a rapidly-administered vaccine plus NPIs (green curve) reduced the case rate in the period encompassed by the red rectangle to 2.01 × 10^−9^ cases/thousand/year, virtually ending the epidemic.

Next, the speed of vaccination and its interaction with duration of immunity warrant some explanation.

### The Importance of Speed of Vaccination

The vaccination speed was implemented by taking a percentage (between 0% and 1%) of the susceptibles and disease-induced immunes per day and adding them to day 1 of the vaccine-induced immunity (viim) compartment. The speed of vaccination had a major effect on the success of the vaccination campaign.

Figure 3 shows the results of an annual vaccination campaign that starts at one year and continues every year thereafter, always with an excess of available vaccine doses. The immunity has a duration of one year. However, this vaccination campaign is unusual because this population never had infectives, and therefore *all immunity is due solely to vaccination*.

**Figure 3.**
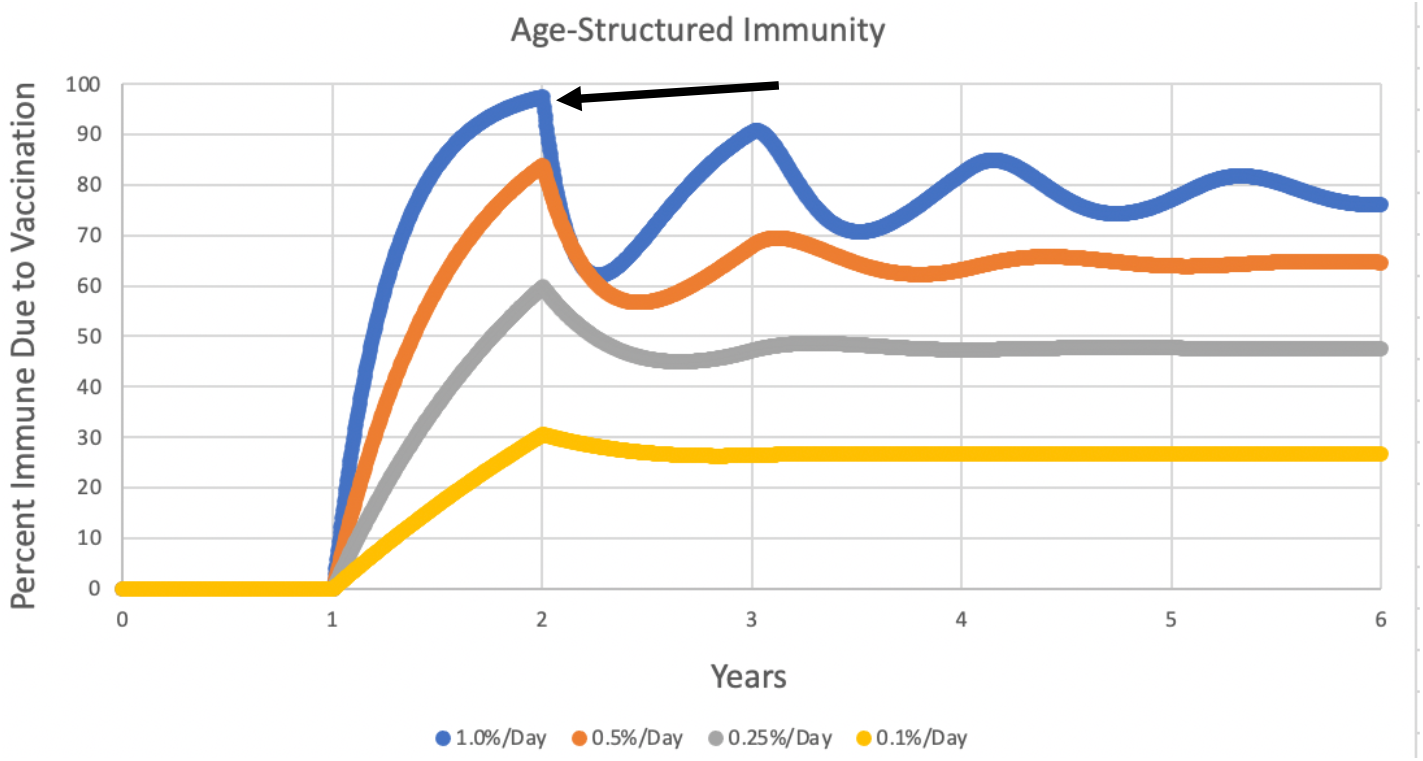
Percent of immune individuals in a population that never had infectives, but begins undergoing vaccination at speeds of 1% (blue), 0.5% (red), 0.25% (gray), or 0.1% (yellow) per day after one year. Vaccine-induced immunity lasted one year. The arrow indicates the point where immunity started to expire in the 1% per day curve.

Note that each curve in Figure 3 rises to a peak, and then experiences a decline one year after vaccination started (arrow in Figure 3). This decline is due to the fact that the first individuals that became vaccinated have “timed out” of immunity and have returned to the susceptibles. Eventually, immunity reaches an equilibrium that depends on the vaccination speed. At low vaccination speeds, the vaccinated portion of the population reaches an equilibrium too low to induce herd immunity because as many individuals are timing out of immunity as are being vaccinated. This means that low vaccination speed can weaken the effectiveness of even an excellent vaccine because the population can only reach herd immunity with substantial help from immunity caused by disease.

Figure 4 was done using the same parameters and the same axes as in Figure 3, but with an exponential rather than an age-structured model of immunity loss. This different immunity model did not change the conclusion--the slower vaccination speeds fall short of an equilibrium that will induce herd immunity by vaccination alone.

**Figure 4.**
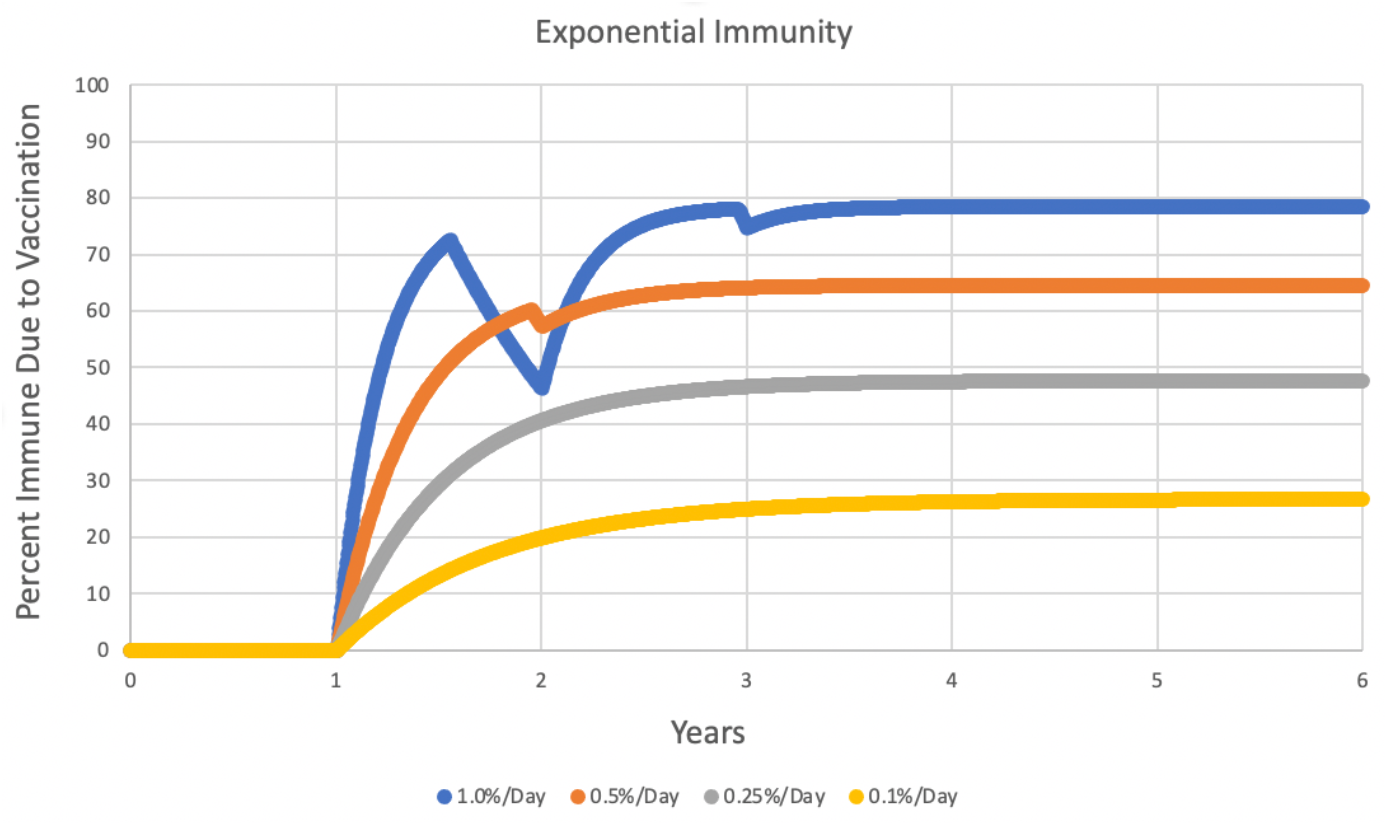
Identical conditions as in Figure 3, but with use of the exponential model of loss of immunity. The sharp decline in the blue curve is caused by the fact that vaccination started at year one but the vaccine supply was exhausted after six months because of the high vaccination rate. This cessation of vaccination caused an immediate decline because individuals are always “leaking” from the immune compartment in exponential immunity. The immunity only rises again when vaccination starts again at the beginning of year 2.

The fact that the curves in Figs. 3 and 4 level off because of expiration of immunity suggests that longer *durations* of immunity might allow even low vaccination speeds to establish herd immunity. Fig. 5 shows the course of immunity in three age-structured simulations that had no infectives and a vaccination speed of 0.25% per day, but durations of immunity of 6 months, 12 months, and 24 months. It is apparent that a longer duration of vaccination-induced immunity increases the ability of moderate vaccination rates to control the disease.

**Fig. 5.**
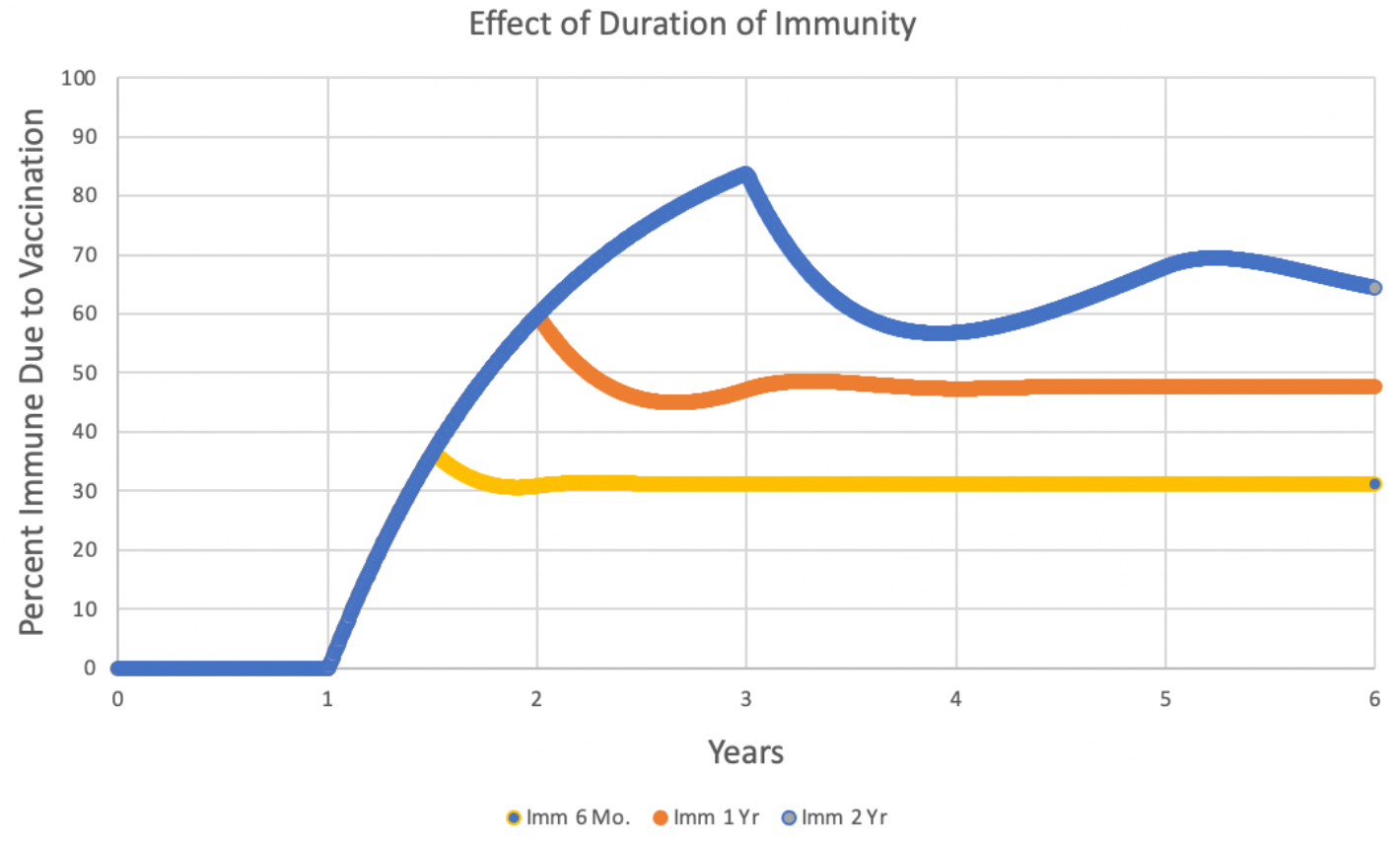
Immunity conferred by vaccination given at 0.25% of the population per day using age-structured immunity. Durations of immunity were 6, 12, and 24 months. All curves followed the blue curve in their early stages.

All the following simulations in this paper used the age-structured model of immunity.

### Case Rates

The main response variable is the number of COVID cases per thousand individuals per year (starting one year after vaccination began). Figure 6 shows the case rates actually experienced by the United States during the first 18 months of the COVID pandemic (*Wikipedia*, 2021). These rates are based on reports of confirmed infections submitted by states, and are probably an underestimate by a factor of almost five (Kalish *et al*., 2021), or by a factor of eight (Ke *et al*., 2021), especially early in the pandemic.

**Figure 6.**
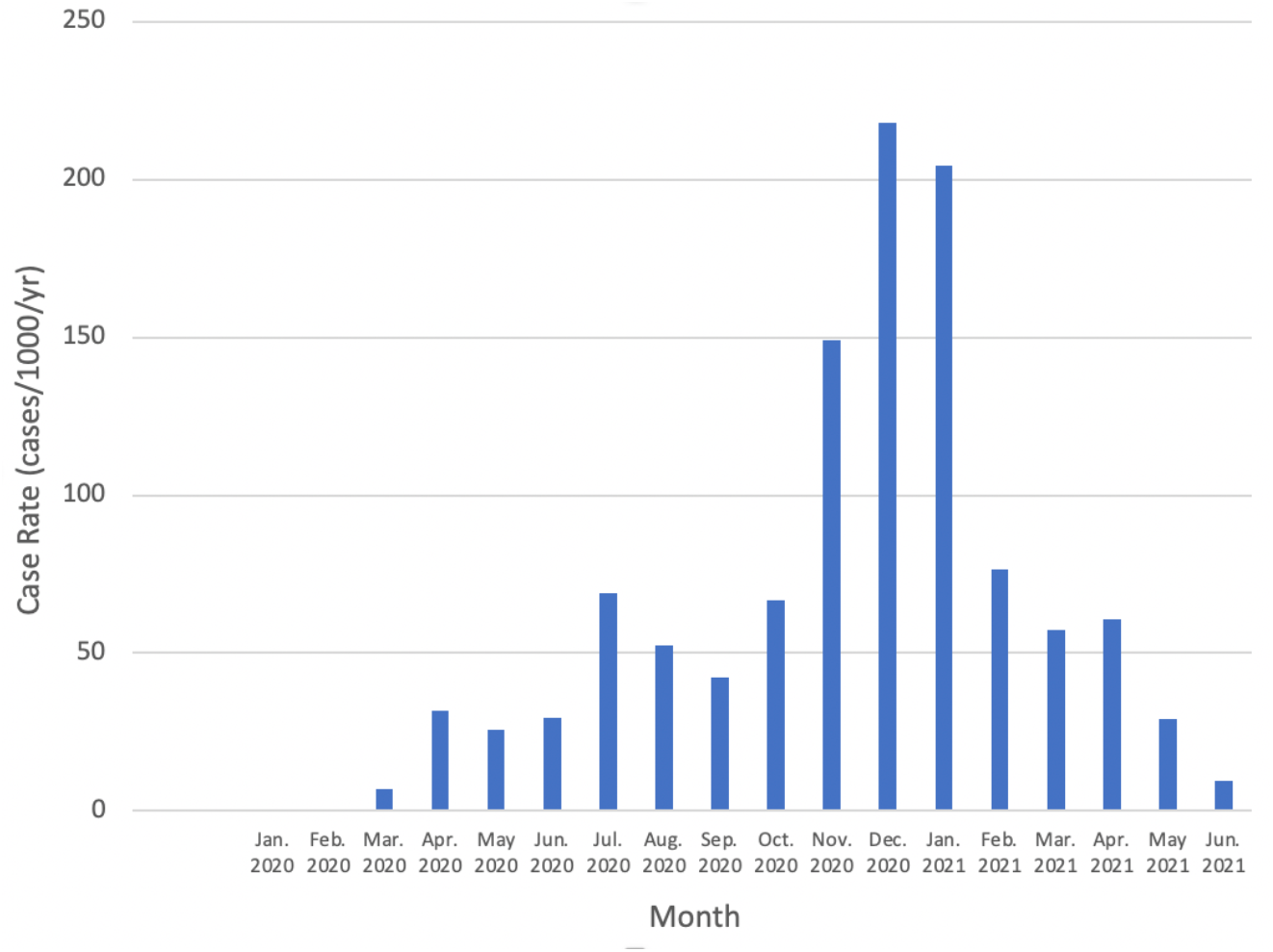
Reported COVID case rates in the United States from January of 2020 through June of 2021, expressed in cases per thousand per year *if the rates experienced in that month were to be continued for an entire year*. The numbers for the first two months were less than 0.001.

Thus, for example, if a case rate in the tables that follow is about 200 cases/thousand/year, this means that the cases for the four-year period after the introduction of the vaccine averaged as high as during the month of January, 2021. A rate that *averaged* that high over four years would be far more damaging than the single month of January, 2021. This must be kept in mind as the results are evaluated.

### Review of the Simulation Scenario

Recall that this paper’s scenario is that a COVID-19 epidemic starts at year zero in Figure 2, and during the first year it is mitigated only by NPIs such as masking and social distancing. Then, at the one-year anniversary of the start of the epidemic, a vaccine is introduced (although in some simulations its rate of distribution will be zero). This vaccine is then distributed for the remainder of the simulation. The main responding variable will be the number of new infections per thousand per year in years 2, 3, 4, and 5, the post-vaccine era, indicated by the red rectangle on Figure 2.

In the first set of simulations (Tables 1-11), the R_0_ of the pathogen remained at 2.3. However, to simulate the appearance of a new, more transmissible variant, a set of simulations in Tables 12-16 increased the R_0_ from 2.3 to 3.0, 4.0, 5.0 or 6.0 between 500 days and 600 days. The R_0_ then remained at this higher value for the rest of the simulation.

**Table 1.** The effect of vaccination at different speeds, with different fractions of the population willing to accept the vaccine. Population 1 accepts the vaccine and Population 2 does not. R_0_ = 2.3, immunity lasts one year and is followed by annual booster vaccinations in cases where vaccination is used. Populations 1 and 2 interact randomly with each other. The variable shown is cases/thousand/year in years 2, 3, 4, and 5 in the combined (1 + 2) population.

| Vac. Rate<br>(%/ Day) | 1 = 100%<br>2 = 0% | 1 = 90%<br>2 = 10% | 1 = 75%<br>2 = 25% | 1 = 50%<br>2 = 50% | 1 = 25%<br>2 = 75% |
| --- | --- | --- | --- | --- | --- |
| 0% | 159.83 |  |  |  |  |
| 0.10% | 13.56 | 17.36 | 25.69 | 51.89 | 97.64 |
| 0.25% | 1.64 | 3.07 | 6.15 | 17.75 | 60.28 |
| 0.50% | $2.06 \times 10^{-9}$ | $1.47 \times 10^{-5}$ | 1.42 | 8.49 | 38.99 |
| 1.00% | 0 | 0 | $3.45 \times 10^{-6}$ | 4.65 | 27.37 |

### Vaccination Speed and Percent of Vaccine-Compliant Individuals

Given that efficacy of the simulated “ideal” vaccine is always 100%, the most important vaccination variables are the speed of vaccination (percent of Population 1 susceptibles and disease-induced immune individuals per day), and the percent of the population that accepts vaccination. Table 1 shows the results for one year of immunity.

Recall that 10 cases/thousand/year could be considered a rough dividing line between unacceptable and acceptable rates of disease over the four post-vaccine years.

### Duration of Vaccine-Induced Immunity

As implied by Fig. 5, the success of the vaccine varied strongly with the duration of immunity:

Since a longer duration of immunity markedly reduces the case rate, could a vaccine that induces *permanent* immunity end the epidemic? Table 3 shows that the answer is “probably not.” While permanent immunity sharply reduces case rates, the outcome depends on the vaccination speed and the mix of Population 1 and Population 2. Since Population 2 is never vaccinated, the epidemic can only be terminated with a fast vaccination speed *and* a small Population 2:

**Table 2.** Post-vaccine case rates for immunity lasting 6, 9, 12, 18 and 24 months for a population that is 75% Population 1 and 25% Population 2. R_0_ = 2.3.

| Vac. Rate<br>(%/ Day) | 6 Months | 9 Months | 12 Months | 18 Months | 24 Months |
| --- | --- | --- | --- | --- | --- |
| 0% | 269.16 | 196.38 | 159.83 | 126.68 | 77.06 |
| 0.10% | 94.04 | 43.59 | 25.69 | 12.58 | 7.31 |
| 0.25% | 20.63 | 10.19 | 6.15 | 2.67 | 0.79 |
| 0.50% | 6.51 | 3.11 | 1.42 | $5.49 \times 10^{-4}$ | $3.20 \times 10^{-5}$ |
| 1.00% | 1.64 | 0.12 | $3.45 \times 10^{-6}$ | $2.60 \times 10^{-10}$ | $1.78 \times 10^{-9}$ |

**Table 3.**
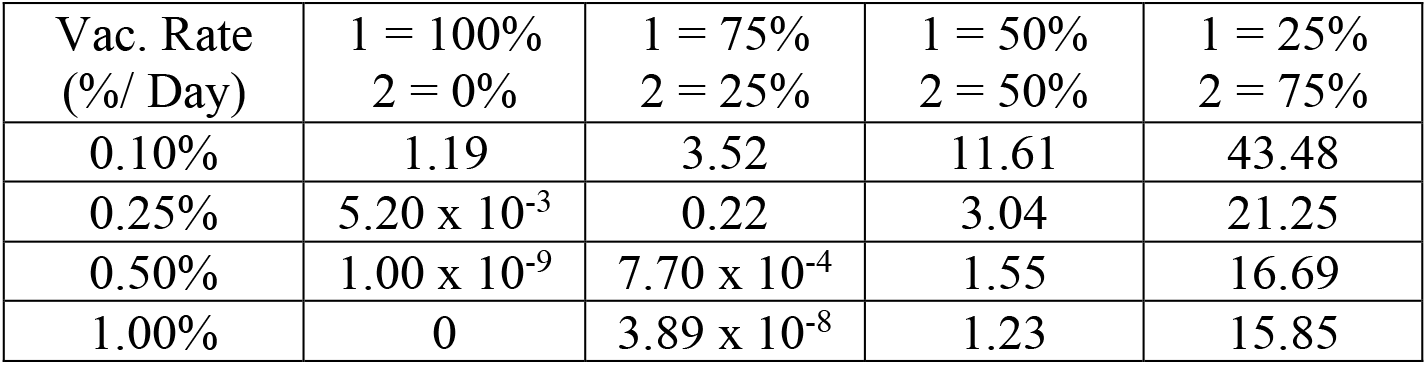
Post-vaccine case rates for disease-induced immunity that lasts one year combined with *permanent* vaccine-induced immunity. R_0_ = 2.3. Compare with Table 1.

| Vac. Rate<br>(%/ Day) | 1 = 100%<br>2 = 0% | 1 = 75%<br>2 = 25% | 1 = 50%<br>2 = 50% | 1 = 25%<br>2 = 75% |
| --- | --- | --- | --- | --- |
| 0.10% | 1.19 | 3.52 | 11.61 | 43.48 |
| 0.25% | $5.20 \times 10^{-3}$ | 0.22 | 3.04 | 21.25 |
| 0.50% | $1.00 \times 10^{-9}$ | $7.70 \times 10^{-4}$ | 1.55 | 16.69 |
| 1.00% | 0 | $3.89 \times 10^{-8}$ | 1.23 | 15.85 |

### Fading Interest in Vaccination

It could be anticipated that some individuals who get the vaccine the first time it is offered might ignore the vaccine in following years. This is called “vaccination fading” below. Note that it is fading of *interest in getting* the vaccine, not a fading of vaccine efficacy. In Table 4, all Population 1 individuals were eligible for vaccination when the vaccine was first offered, but after that, the doses available to Population 1 were reduced by 25%, 50%, or 75% per year.

**Table 4.** Vaccine era case rates for several combinations of Populations 1 and 2 (columns) subjected to several degrees of vaccination fading (rows). Recall that Population 1 is vaccinated and Population 2 is not. Immunity lasted one year, R_0_ = 2.3, and vaccination rate was 0.5% of the Population 1 susceptibles and disease-induced immunes per day.

| Vac. Fade<br>(%/Year) | 1 = 100%<br>2 = 0% | 1 = 90%<br>2 = 10% | 1 = 75%<br>2 = 25% | 1 = 50%<br>2 = 50% | 1 = 25%<br>2 = 75% |
| --- | --- | --- | --- | --- | --- |
| 0% | $2.01 \times 10^{-9}$ | $1.47 \times 10^{-5}$ | 1.42 | 8.49 | 38.99 |
| 25% | $2.01 \times 10^{-9}$ | $7.15 \times 10^{-5}$ | 2.13 | 9.89 | 42.21 |
| 50% | 32.22 | 41.44 | 48.62 | 61.60 | 88.26 |
| 75% | 73.20 | 88.01 | 97.55 | 108.67 | 122.33 |

These reductions built on one another, so that with 50% reduction/year, the number of doses available to Population 1 would be 100% of those needed in the first vaccine year, 50% in the second year, 25% in the third year, and 12.5% in the fourth year. Once vaccination fading got over 25% per year, it could lead to large increases in the vaccine era case rate:

For comparison, if there was no vaccination at all, the case rate for one year of immunity would be 159.83 cases/thousand/year for one year of immunity for both Populations 1 and 2.

The results in Table 4 are for one year of immunity. Table 5 shows that longer durations of immunity could still maintain low case rates even when fading of interest in vaccination was severe:

**Table 5.** Post-vaccine era case rates for a population that was 75% Population 1 and 25% Population 2. R_0_ = 2.3, vaccination rate was 0.5%/day. Immunity lasted from 6 to 24 months.

| Vac. Fade<br>(%/Year) | 6 Months | 9 Months | 12 Months | 18 Months | 24 Months |
| --- | --- | --- | --- | --- | --- |
| 0% | 6.52 | 3.11 | 1.42 | $5.49 \times 10^{-4}$ | $3.20 \times 10^{-5}$ |
| 25% | 6.52 | 3.11 | 2.13 | $5.49 \times 10^{-4}$ | $3.20 \times 10^{-5}$ |
| 50% | 40.17 | 50.71 | 48.62 | 9.62 | $3.20 \times 10^{-5}$ |
| 75% | 128.12 | 117.34 | 97.55 | 62.81 | 19.07 |

### Abandoning NPIs

Recall that in nearly all simulations above, the vaccines were combined with NPIs that produced a maximum lockdown parameter (lkd) of 0.5 when the rate of infection was high. This reduced R_0_ by a factor of 2 and resulted in much lower rates of infection when the R_0_ was 2.3, as in all simulations so far. However, as vaccination becomes more common and rates of disease drop, there might be a tendency for the public to stop masking and social-distancing because they think that vaccination has replaced NPIs. Table 6 simulates an epidemic that introduces a vaccine on day 366 and then drops its lockdown parameter from 0.5 to either 0.25 or 0.0 starting on day 500. This drop happens in both Population 1 and Population 2. Vaccination continues in Population 1 after day 500. The effects of abandoning NPIs were powerful, even though vaccination continued.

**Table 6.** The effect of reducing NPIs starting on day 500 on vaccine-era case rates in cases per thousand per year. Duration of immunity is one year, and R_0_ = 2.3. The population consisted of 75% Population 1 (seeks vaccination) and 25% Population 2 (refuses vaccines). Interest in getting a vaccine does not fade with time.

| Lkd after 500 days | 0%/Day | 0.1%/Day | 0.25%/Day | 0.5%/Day | 1%/Day |
| --- | --- | --- | --- | --- | --- |
| 0.5 | 159.83 | 25.69 | 6.15 | 1.42 | $3.45 \times 10^{-6}$ |
| 0.25 | 363.47 | 247.77 | 87.24 | 4.59 | $4.13 \times 10^{-6}$ |
| 0 | 442.52 | 312.51 | 256.41 | 96.00 | $5.06 \times 10^{-6}$ |

#### Vaccination Speed and Weakening of NPIs

A major conclusion of Table 1 was that faster vaccination rates resulted in greater reduction in case rates. Table 6 shows that faster vaccination rates also can avert damage done by stopping NPIs, but also predicts the weakening of NPIs will cause remarkably higher case rates:

The remarkable increase in case rates as NPIs decline is the major finding. Note the explosion of case rates for a moderate (and realistic) vaccination rate like 0.25%/day. However, also note how the rightmost column of Table 6 shows not only very low case rates, but also little effect of dropping NPIs to zero after 500 days.

#### Percent of Population Accepting Vaccination and Weakening of NPIs

Table 6 shows results for 75% Population 1 and 25% Population 2. Abandoning NPIs causes worse consequences when a high percentage of the population (Population 2) doesn’t seek vaccination. Table 7 shows that if the whole population is getting vaccinated (Population 2 is zero), the decision to abandon NPIs has only a minor effect on case rates. However, if there is a significant Population 2, cases that start in Population 2 increase markedly as NPIs are abandoned.

**Table 7.** The interaction between abandoning of NPIs and the percent of the population that does not seek vaccination. R_0_ = 2.3, vaccination rate of Population 1 is 0.5%/day; immunity lasts one year; interest in getting vaccinated by Population 1 does not fade with time. At 500 days into the simulation, both Population 1 and 2 reduce their lockdown parameter from 0.5 to either 0.25 or 0.0. The columns show different mixes of Population 1 and Population 2.

| Lkd after 500 days | 1 = 100%<br>2 = 0% | 1 = 90%<br>2 = 10% | 1 = 75%<br>2 = 25% | 1 = 50%<br>2 = 50% | 1 = 25%<br>2 = 75% |
| --- | --- | --- | --- | --- | --- |
| 0.5 | $2.01 \times 10^{-9}$ | $1.47 \times 10^{-5}$ | 1.42 | 8.49 | 38.99 |
| 0.25 | $2.88 \times 10^{-9}$ | $2.41 \times 10^{-5}$ | 4.59 | 124.91 | 315.43 |
| 0 | $4.62 \times 10^{-9}$ | $5.09 \times 10^{-5}$ | 96.00 | 300.85 | 333.18 |

The harmful effect of abandoning NPIs can be seen in Figure 7, which shows the course of the infection for the top and bottom cases in the 75%/25% column in Table 7.

**Figures 7a and 7b.**
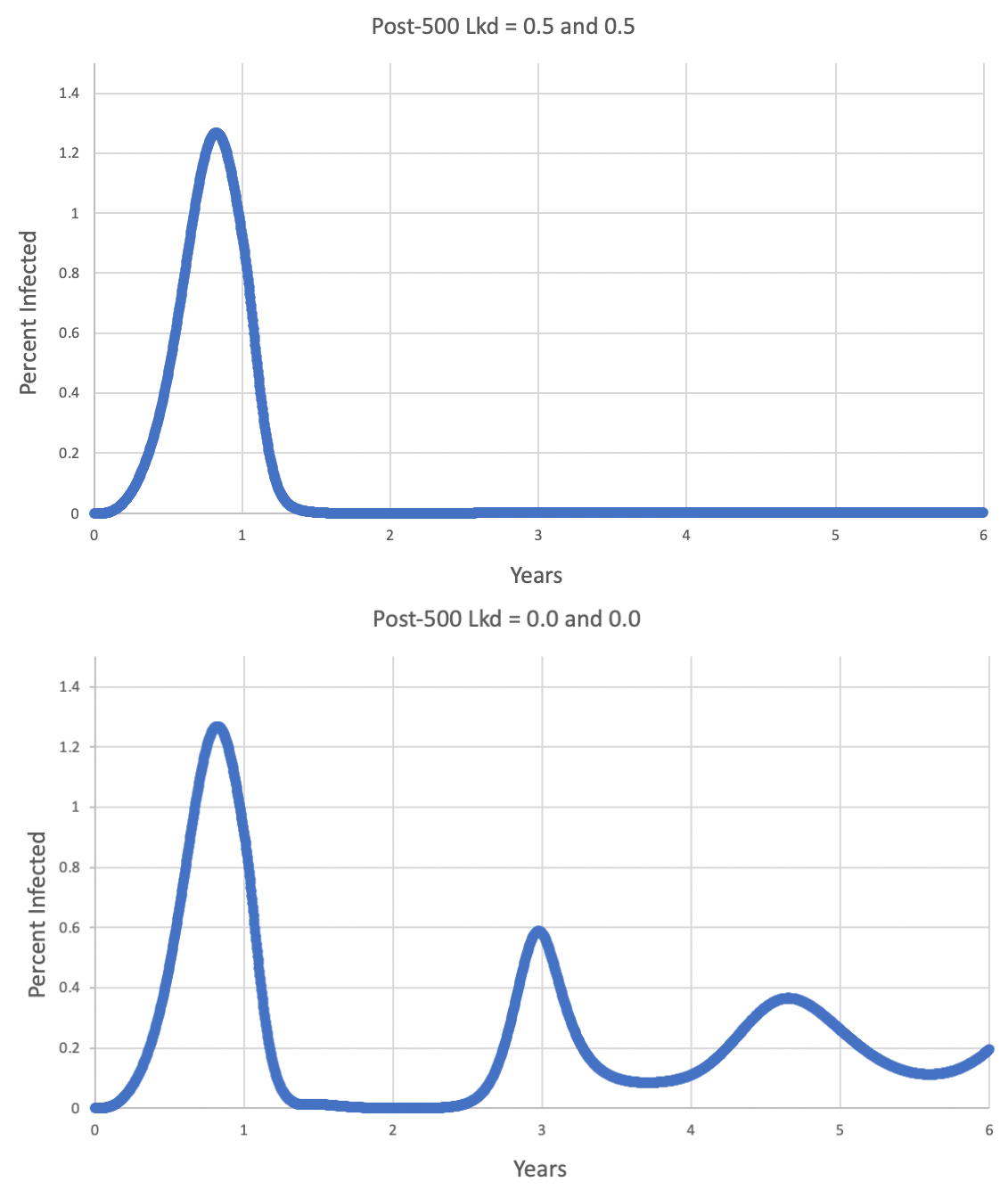
Course of epidemic in a population that is 75% Population 1 and 25% Population 2, with one year of immunity. A change in NPIs may occur at 500 days (1.37 years). In Fig. 7a (above), both populations maintain a maximum lockdown parameter of 0.5 past 500 days. In Fig. 7b (below), both populations drop their lockdown parameter to zero after 500 days. In both cases, vaccination continues annually in Population 1. The post-vaccine case rate is 1.42 cases/thousand/year in Fig. 7a and 96.00 in Fig. 7b.

Table 7 shows that when NPIs are weakened, case rates increase as the size of Population 2 rises. Table 8 looks at this from another viewpoint. As the size of Population 2 increases, what vaccination speeds in Population 1 are necessary to keep Population 1’s case rate at 10 cases/thousand/year? Note that this case rate is for Population 1 only.

**Table 8.**
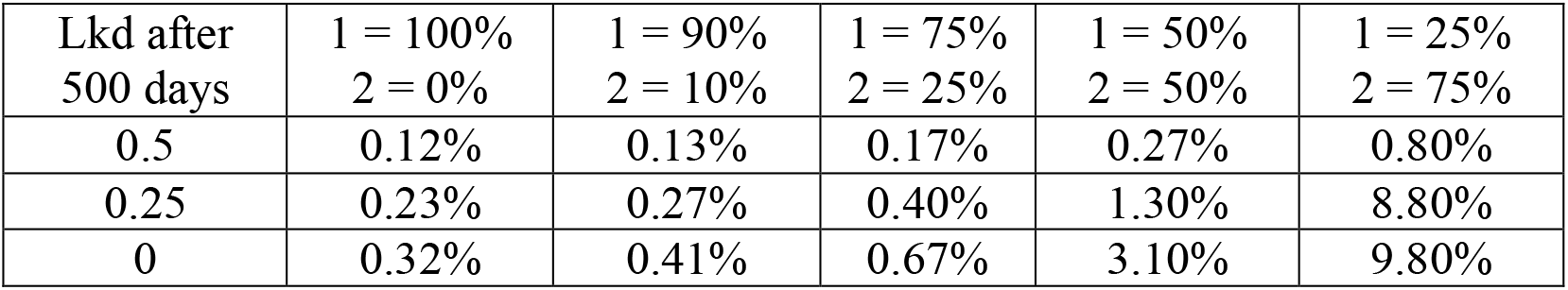
The effect of the size of Population 2 on the vaccination speed (% per day) necessary to give Population 1 a case rate of approximately 10 cases/thousand/year. R_0_ = 2.3, immunity lasts one year; interest in getting vaccinated by Population 1 does not fade with time. At 500 days into the simulation, both Population 1 and 2 either keep their lockdown parameter at 0.5 or change it to 0.25 or 0.0.

#### Duration of Immunity and Weakening of NPIs

The duration of immunity has a powerful effect if NPIs are abandoned. Longer durations of immunity protect the population from NPI reduction:

### Social Isolation of Population 1 from Population 2

All the simulations above assumed that Population 1 and Population 2 mixed randomly, and that the probability that a Population 1 individual would interact with a Population 2 individual was proportional to their relative abundances. However, vaccination is more common in some US states and social groups than in others. This suggests that Populations 1 and 2 might be partially isolated from one another. Understanding the consequences will require considering the case rates of Population 1 and Population 2 separately, not just the rate for the combination population:

Willingness to maintain NPIs after the vaccine arrived was an important contributor to success in Tables 6-9. As NPIs are abandoned, Population 2 suffers from high rates of disease. Table 11 (especially the rightmost column) shows how vaccinated Population 1 can benefit from reduced interaction with Population 2, especially when NPIs are abandoned.

**Table 9.** Comparison of post-vaccine case rates in simulations with 6 to 24 months of immunity. In all cases, R_0_ = 2.3, the vaccination rate is 0.5% of the susceptibles and disease-induced immunes/day, interest in vaccinations does not fade, and 75% of the population is willing to accept vaccination. At 500 days into the simulation, both Population 1 and 2 change their maximum lockdown parameter from 0.5 to either 0.5, 0.25, or 0.0.

| Lkd after<br>500 days | 6 Months | 9 Months | 12 Months | 18 Months | 24 Months |
| --- | --- | --- | --- | --- | --- |
| 0.5 | 6.52 | 3.11 | 1.42 | $5.49 \times 10^{-4}$ | $3.20 \times 10^{-5}$ |
| 0.25 | 165.19 | 21.04 | 4.59 | $1.10 \times 10^{-3}$ | $6.54 \times 10^{-5}$ |
| 0 | 437.45 | 226.24 | 96.00 | $4.34 \times 10^{-3}$ | $2.67 \times 10^{-4}$ |

**Table 10.** Post-vaccine era case rates in Populations 1 and 2 with various degrees of interaction. An interaction parameter of 1.0 indicates random mixing between the populations; an interaction parameter of 0.0 indicates no interaction between the populations. R_0_ = 2.3, immunity lasts one year, the mix of Populations 1 and 2 is 75%/25%, the vaccination rate is 0.5%/day, and interest in vaccines does not fade from year to year. Both populations maintain their maximum lockdown parameters at 0.5 after 500 days.

| Interaction | Pop. 1 | Pop. 2 | Combined |
| --- | --- | --- | --- |
| 1.0 | 0.98 | 2.74 | 1.42 |
| 0.5 | 1.05 | 4.61 | 1.94 |
| 0.1 | 1.73 | 57.51 | 15.67 |
| 0.0 | $2.06 \times 10^{-9}$ | 159.17 | 39.79 |

**Table 11.** Post-vaccine case rates, 75% Population 1 and 25% Population 2, R_0_ = 2.3, 12 months of immunity, vaccination rate of 0.5%/day in Population 1, no fading of interest in getting vaccinated, maximum lockdown parameters after 500 days of 0.5 and 0.0, with several degrees of interaction between Population 1 and Population 2.

| Interac. | Pop. | Lkd = 0.5 | Lkd = 0 |
| --- | --- | --- | --- |
| 1.0 | 1 | 0.98 | 68.14 |
|  | 2 | 2.74 | 179.58 |
| 0.5 | 1 | 1.05 | 64.22 |
|  | 2 | 4.61 | 222.64 |
| 0.1 | 1 | 1.73 | 29.59 |
|  | 2 | 57.51 | 379.77 |
| 0.0 | 1 | $2.06 \times 10^{-9}$ | $4.68 \times 10^{-9}$ |
|  | 2 | 159.17 | 442.59 |

In the lowest, rightmost cell of Table 11, the highest percent of infection seen in Population 1 during the four post-vaccine years was 2.6 × 10^−9^ %; in Population 2, it was 25%.

### Evolution of a More Transmissible Variant

Tables 1-11 assumed that the pathogen retained its “2020” R_0_ of 2.3. However, in 2021, variants that are more transmissible have risen to prominence. This was simulated by increasing R_0_ from its original value of 2.3 to a higher target R_0_ between 500 and 600 days into the simulation. Even small increases in R_0_ had a pronounced effect on the post-vaccine case rates (Table 12):

**Table 12.** Ability of several vaccination rates to control case rates as the base R_0_ of 2.3 was increased to various final R_0_s between 500 and 600 days of the simulation. Duration of immunity one year, 75% Population 1 and 25% Population 2, no fading of interest in getting the vaccine, lockdown parameter maintained at 0.5 after 500 days, random interaction between Population 1 and Population 2. Column headings show the final R_0_ from 600 days to the end of the simulation.

| Vac. Rate per Day | $R_0$ 2.3 | $R_0$ 3.0 | $R_0$ 4.0 | $R_0$ 5.0 | $R_0$ 6.0 |
| --- | --- | --- | --- | --- | --- |
| 0% | 159.83 | 370.50 | 604.54 | 457.70 | 481.33 |
| 0.1% | 25.69 | 177.51 | 394.47 | 492.32 | 543.84 |
| 0.25% | 6.15 | 37.21 | 221.40 | 344.84 | 389.16 |
| 0.5% | 1.42 | 8.29 | 68.60 | 219.31 | 290.40 |
| 1.0% | $3.45 \times 10^{-6}$ | 1.79 | 13.78 | 77.62 | 186.29 |

Although increasing the pathogen R_0_ always caused an increase in case rate, longer durations of immunity reduced the damage done by more transmissible variants:

Another way of countering more transmissible variants is to reduce the size of the unvaccinated Population 2. Vaccination of almost the whole population can be effective, but if more than 25% of the population refuses vaccination, an aggressive new variant drives case rates very high:

As in Table 8, in addition to presenting case rates, it is also instructive to present the vaccination speed that is necessary to maintain a case rate of 10 cases/thousand/year in Population 1:

Tables 6-9 showed that the weakening of NPIs after a vaccine is implemented could cause an upsurge in the case rate even if vaccination of Population 1 was continuing and even if the R_0_ of the pathogen remained at the original value of 2.3. A new, more transmissible variant would worsen the consequences of reducing NPIs. On the other hand, if NPIs can be *strengthened* when new variants threaten, the transmissibility of the variants might be lessened. In Table 16, the maximum lockdown parameter remained at 0.5 up until 500 days, and then either decreased or increased to the values in the first column of the table. This decrease or increase in NPIs occurred in both Population 1 and Population 2. Vaccination continued in Population 1.

**Table 13.** A population consisting of 75% Population 1 and 25% Population 2, with a daily vaccination rate of 0.5%/day. Other characteristics are in the heading for Table 12.

| Duration (Months) | $R_0$ 2.3 | $R_0$ 3.0 | $R_0$ 4.0 | $R_0$ 5.0 | $R_0$ 6.0 |
| --- | --- | --- | --- | --- | --- |
| 6 | 6.52 | 47.98 | 365.49 | 565.34 | 698.93 |
| 9 | 3.11 | 15.35 | 150.09 | 350.67 | 389.25 |
| 12 | 1.42 | 8.29 | 68.60 | 219.31 | 290.40 |
| 18 | $5.49 \times 10^{-4}$ | 3.47 | 29.95 | 125.14 | 182.16 |
| 24 | $3.20 \times 10^{-5}$ | 0.90 | 18.24 | 76.12 | 152.92 |

**Table 14.** A population consisting of various percentages of Population 1 with one year of immunity. Vaccination rate = 0.5%/day. Other characteristics are in the heading for Table 12.

| % Pop. 1 | $R_0$ 2.3 | $R_0$ 3.0 | $R_0$ 4.0 | $R_0$ 5.0 | $R_0$ 6.0 |
| --- | --- | --- | --- | --- | --- |
| 100% | $2.00 \times 10^{-9}$ | $5.61 \times 10^{-2}$ | 4.68 | 23.13 | 96.66 |
| 90% | $7.77 \times 10^{-4}$ | 2.41 | 14.28 | 86.47 | 194.58 |
| 75% | 1.42 | 8.29 | 68.60 | 219.31 | 290.40 |
| 50% | 8.49 | 59.60 | 273.48 | 344.08 | 420.07 |

**Table 15.** The speed of vaccination (%/day) necessary to maintain a case rate of 10 cases/thousand/year in Population 1 when total population includes various percentages of Population 1. One year of immunity, lockdown parameter is maintained at 0.5, random interaction between Populations 1 and 2.

| % Pop. 1 | R<sub>0</sub> 2.3 | R<sub>0</sub> 3.0 | R<sub>0</sub> 4.0 | R<sub>0</sub> 5.0 | R<sub>0</sub> 6.0 |
| --- | --- | --- | --- | --- | --- |
| 100% | 0.12% | 0.24% | 0.41% | 0.59% | 0.74% |
| 90% | 0.13% | 0.28% | 0.53% | 0.81% | 1.15% |
| 75% | 0.17% | 0.39% | 0.90% | 1.80% | 3.40% |
| 50% | 0.58% | 1.00% | 4.10% | 12.20% | 25.00% |

**Table 16.** A population with one year of immunity consisting of 75% Population 1 and 25% Population 2 with vaccination speed at 0.5%/day. Random interaction between Populations 1 and 2. At 500 days, the lockdown parameter changed from 0.5 to the values shown. Between 500 and 600 days, the R_0_s of the pathogen increased from 2.3 to the values shown in the column headings.

| Lkd. after 500 Days | R<sub>0</sub> 2.3 | R<sub>0</sub> 3.0 | R<sub>0</sub> 4.0 | R<sub>0</sub> 5.0 | R<sub>0</sub> 6.0 |
| --- | --- | --- | --- | --- | --- |
| 0.0 | 96.00 | 284.67 | 408.10 | 415.35 | 499.60 |
| 0.25 | 4.58 | 114.21 | 292.33 | 402.71 | 422.84 |
| 0.5 | 1.42 | 8.29 | 68.60 | 219.31 | 290.40 |
| 0.6 | 0.91 | 5.05 | 19.56 | 81.42 | 199.45 |
| 0.7 | 0.99 | 3.62 | 9.79 | 22.62 | 55.03 |
| 0.8 | 0.77 | 2.82 | 6.46 | 11.48 | 18.77 |

### An “Optimistic” and a “Pessimistic” Scenario

The tables above presented many options for the post-vaccine case rates, but tended to vary one variable at a time and hold the others at default values. In order to focus on a reduced number of outcomes, Table 17 presents an “optimistic” and a “pessimistic” set of parameters and the model’s prediction of the case rates in the post-vaccine era.

**Table 17.**
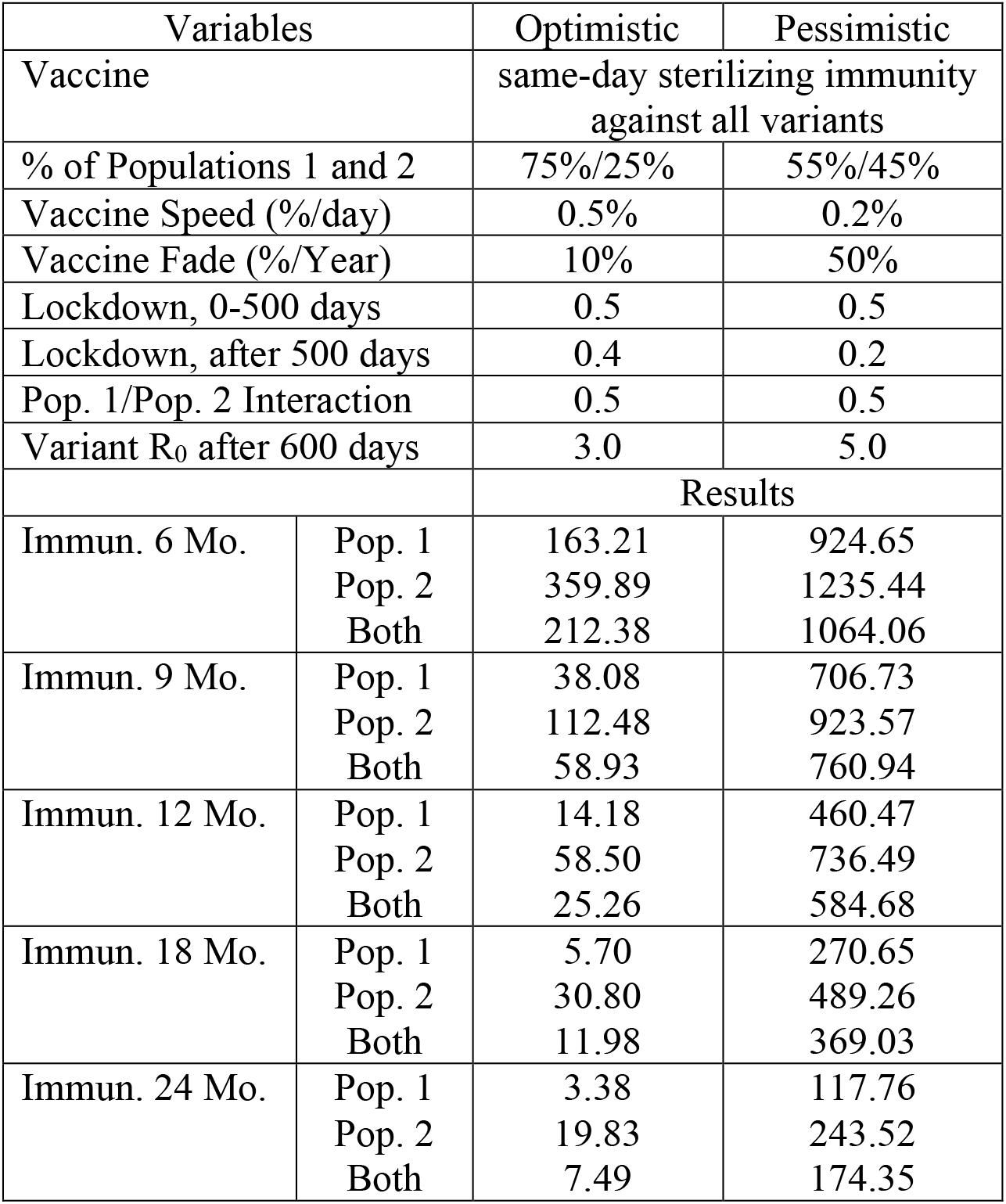
Key parameters for an “optimistic” and a “pessimistic” COVID-19 future, and the post-vaccine case rates they would produce, in cases/thousand/year. “Vac. Fade” represents loss of interest in getting annual booster shots after the first vaccine is received. “Both” indicates the case rates of the weighted combination of Populations 1 and 2.

| Variables |  | Optimistic | Pessimistic |
| --- | --- | --- | --- |
| Vaccine |  | same-day sterilizing immunity against all variants |  |
| % of Populations 1 and 2 |  | 75%/25% | 55%/45% |
| Vaccine Speed (%/day) |  | 0.5% | 0.2% |
| Vaccine Fade (%/Year) |  | 10% | 50% |
| Lockdown, 0-500 days |  | 0.5 | 0.5 |
| Lockdown, after 500 days |  | 0.4 | 0.2 |
| Pop. 1/Pop. 2 Interaction |  | 0.5 | 0.5 |
| Variant $R_0$ after 600 days | | 3.0 | 5.0 |
|  |  | Results |  |
| Immun. 6 Mo. | Pop. 1 | 163.21 | 924.65 |
|  | Pop. 2 | 359.89 | 1235.44 |
|  | Both | 212.38 | 1064.06 |
| Immun. 9 Mo. | Pop. 1 | 38.08 | 706.73 |
|  | Pop. 2 | 112.48 | 923.57 |
|  | Both | 58.93 | 760.94 |
| Immun. 12 Mo. | Pop. 1 | 14.18 | 460.47 |
|  | Pop. 2 | 58.50 | 736.49 |
|  | Both | 25.26 | 584.68 |
| Immun. 18 Mo. | Pop. 1 | 5.70 | 270.65 |
|  | Pop. 2 | 30.80 | 489.26 |
|  | Both | 11.98 | 369.03 |
| Immun. 24 Mo. | Pop. 1 | 3.38 | 117.76 |
|  | Pop. 2 | 19.83 | 243.52 |
|  | Both | 7.49 | 174.35 |

For 12 months of immunity, the post-vaccine case rate in the optimistic simulation was the result of a long period of consistently low rates of disease (Fig. 8); in the pessimistic case, it resulted from several spectacular outbreaks from low levels of disease (Fig. 9). Note the fact that the “optimistic” maximum percent infected is 1.3%, but the “pessimistic” maximum is 35%.

**Fig. 8.**
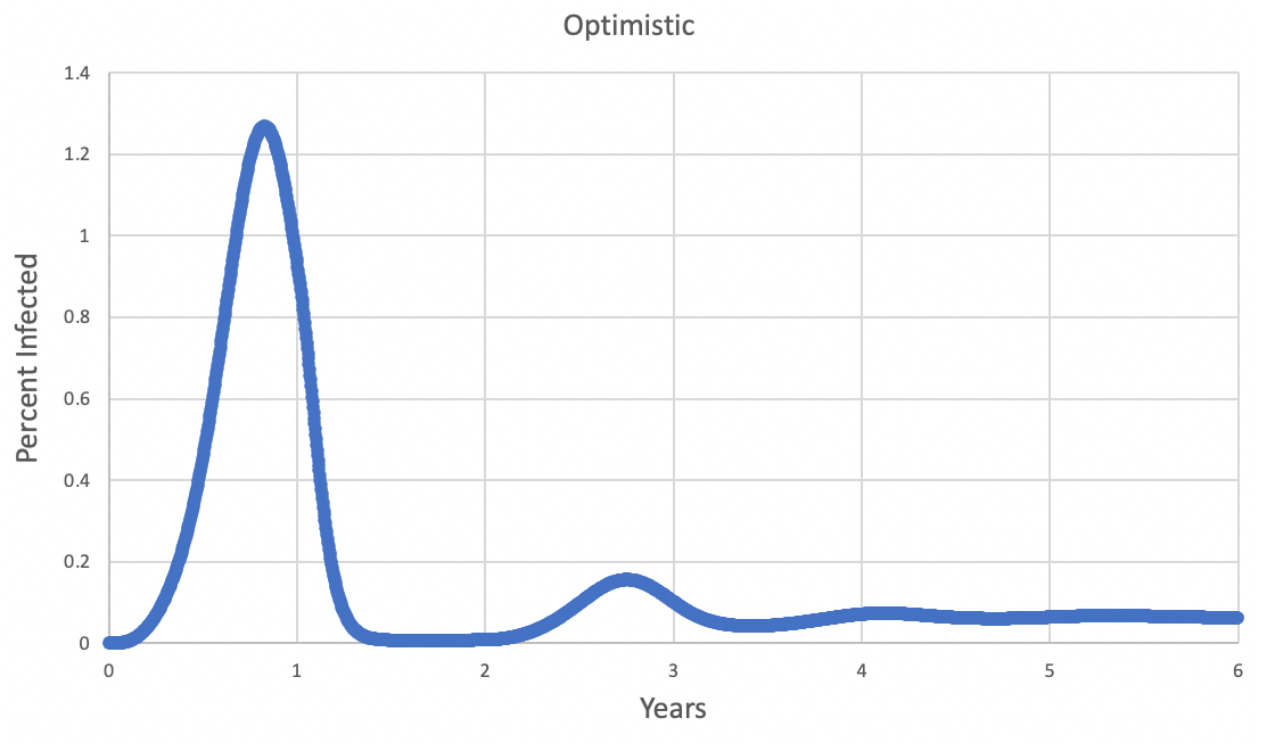
Infection rate in the “optimistic” case for one year of immunity. For the first 1.37 years, the R_0_ was 2.3, and then increased to 3.0 by 1.64 years. Other “optimistic” parameters are in Table 17.

**Fig. 9.**
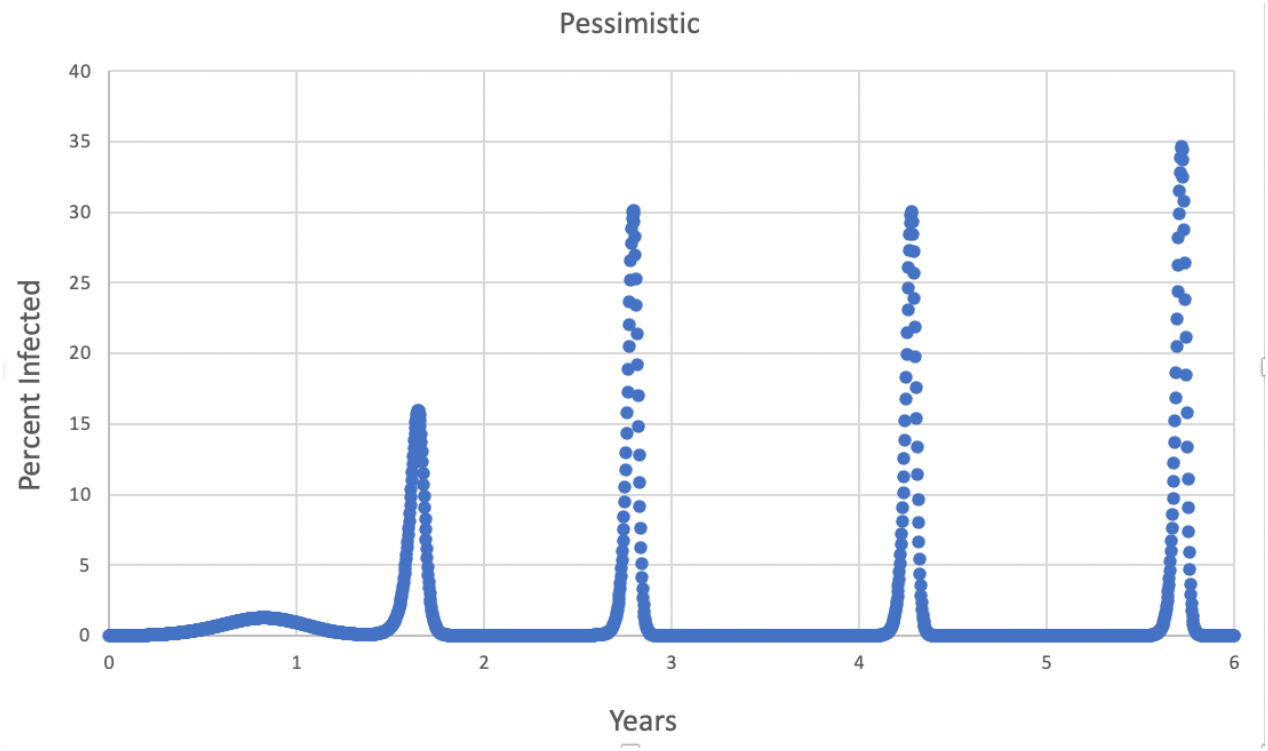
Infection rate in the “pessimistic” case for one year of immunity. For the first 1.37 years, the R_0_ was 2.3, and then increased to 5.0 by 1.64 years. The sharp peaks were driven by expiration of immunity. The small “bump” just before one year is the same size as the large peak in Fig. 8.

## Discussion

Despite the power of vaccines, these simulations disclosed that common situations can impair vaccine success, as measured by COVID case rates in the years following vaccine introduction. The most noteworthy results involved the speed of vaccine administration, the duration of immunity, maintenance of NPIs such as masking even after vaccines are in use, and the development of more transmissible variants.

The speed of vaccination is used many times in this report. For context, the Introduction quoted data from the CDC COVID Data Tracker showing that the exponential decline of the incompletely vaccinated portion of the US 12+ population averaged 0.34%/day from 14 December 2020 to 17 November 2021, and 0.26%/day over the same period for the whole population.

Table 1 shows that if 90-100% of the population can be vaccinated at 0.5% of the susceptibles per day or faster, an epidemic with an R_0_ of 2.3 and vaccine-induced immunity of one year can be essentially ended. The same is true if 25% of the population refuses vaccination, but then the vaccination speed must be 1% per day. Greater unvaccinated portions of the population preclude ending the epidemic, but fast vaccination rates may still keep the case rate below 10 cases/thousand/year.

Table 2 shows the powerful effects of duration of immunity. If immunity lasts only a short time (say, 6 months), there won’t be enough time to vaccinate a large portion of the population before the immunity begins to expire. A slow vaccination rate further reduces the immunity that can be built up before immunity expires. On the other hand, if immunity is long-lasting, even a moderate vaccination rate may be enough to maintain substantial vaccine-induced immunity (Fig. 5). Table 1 showed that while a larger portion of individuals refusing vaccination increases case rates, Table 2 shows that longer duration of immunity reduces case rates. The epidemic can nearly be ended by a vaccination rate of 0.5%/day if immunity lasts two years, but this would require 1%/day with one year of immunity, and the epidemic cannot be ended using realistic vaccination rates with 6 months of immunity.

Table 3 explores this duration question further with a hypothetical vaccine that induces permanent vaccinal immunity. If every member of the population could be vaccinated, this new vaccine could end the epidemic, especially if the vaccination speed is fast. However, Population 2 is beyond the reach of the vaccine. If Population 2 is 50% or more of the total, the epidemic persists at a low level in the unvaccinated population even when vaccine immunity is permanent.

Of course, progressive loss of interest in getting the vaccine (“vaccine fading”) increases case rates in the post-vaccine era (Table 4), but as long as the decline per year is less than 25%, the effect on case rates is small in the first five years after vaccine introduction. Higher rates of vaccine fading cause a rapid escalation of case rates. Table 2 made the point that long durations of immunity allowed slow vaccination rates to be more successful; Table 5 shows that long durations also protect against declining interest in immunizations. As long as the interest in getting a vaccination declines at less than 50% per year, 24 months of immunity produces lower case rates than any shorter duration ever produces, even with no decline in vaccination interest.

One of this paper’s major points is that dropping of NPIs such as masking after vaccinations begin can drastically increase long-term case rates (Table 6). This conclusion has also been reached by most of the papers cited in the Introduction. For their first 499 days, the simulations had a lockdown parameter of 0.5, which cut the realized R_0_ from 2.3 to 1.15 once the infection became common. However, on day 500, some simulations kept the lockdown factor at 0.5, some reduced it to 0.25, and some reduced it to zero. This simulated the CDC announcement on 13 May 2021 that vaccinated individuals could go without masks indoors and no longer needed to social-distance.

The higher case rates as NPIs weaken are due to high case rates in the unvaccinated population. Consider the 0.5%/day column in Table 6. The last line in that table shows a case rate of 96.00 cases/thousand/year when the lockdown parameter goes to zero after 500 days. This 96.00 is due to the averaging of a case rate of 68.14 for Population 1 (75%) and 179.58 for Population 2 (25%). However, if Population 2 is set to zero and all other parameters remain the same for Population 1, the case rate for Population 1 is 4.68 × 10^−9^ cases/thousand/year. The difference between 4.68 × 10^−9^ and 68.14 cases/thousand/year shows that almost all of Population 1’s cases in Table 6 are coming from Population 2.

Table 7 shows the powerful interaction between the size of the unvaccinated population and abandoning NPIs. Where Population 2 is zero (first column), every member of the population can be vaccinated. Population 1 can abandon NPI precautions and still enjoy very low case rates. This is still true when Population 1 is 90%. However, where Population 1 is 50% or lower, this situation rapidly reverses. Case rates reach well over 100 cases/thousand/year, as bad as the worst months of the pandemic in the US. This is presented graphically in Figs. 7a and 7b. The dramatic difference between Figs. 7a and 7b is due to infections that originate in Population 2 when its lockdown factor drops to zero.

Table 8 also investigates the interaction between vaccination speed and the size of Population 2 as NPIs are abandoned. Because vaccination lowers case rates and a greater size of the unvaccinated population raises them, what vaccination speed in Population 1 would be necessary to keep Population 1’s case rate at 10 cases/thousand/year as Population 2 increases in size? When Population 2 is small, the required vaccination rates in Population 1 are attainable, less than 0.5% of the population/day. However, once Population 2 exceeds 25% of the total, the required vaccination rates exceed 1% per day, and nearly exceed 10%/day. These rates would be impossible to sustain on a national scale. The conclusion is clear--NPIs and vaccination must work together. If NPIs are maintained (first line of Table 8), vaccination has a manageable task. However, if NPIs are abandoned, even the most aggressive vaccination effort cannot keep case rates low. It is worth noting that Table 8 uses an R_0_ of 2.3; higher R_0_s (Tables 12-15) also make the task of vaccines more difficult.

Long durations of immunity can mitigate the effect of the slow rate of vaccination (Table 2) because the longer the period of immunity is, the more time there is to complete the vaccination of the population. Table 9 shows that long durations can also blunt the effect of abandoning NPIs. In a population consisting of 75% Population 1 and 25% Population 2, the case rate when both populations abandoned NPIs completely went from 437.45 cases/thousand/year with six months of immunity to 2.67 × 10^−4^ with 24 months of immunity. Case rates for 18 months of immunity were remarkably lower than case rates with 12 months of immunity.

Isolating Population 1 (accepts vaccines) from Population 2 (refuses vaccines) in Table 10 (one year of immunity) tends to exaggerate the case rate difference between Populations 1 and 2. When mixing between Populations 1 and 2 is random (interaction parameter = 1.0), Population 1’s case rate is raised by the presence of Population 2, and Population 2’s case rate is reduced by the presence of Population 1, but both rates are moderate. As the two populations become more isolated from one another, Population 1’s case rate increases slightly but Population 2’s case rate rises quickly, and this causes the combined case rate to increase markedly. When the two populations are completely isolated from one another (interaction parameter = 0), Population 1’s case rate may go to zero or close to it, but Population 2’s case rate is about 150 cases/thousand/year, near the case rate seen in the US in November 2020. Again, this points to Population 2 as the source of Population 1’s disease. Population 2 is suffering from “an epidemic of the unvaccinated,” and being cut off from heavily-infected Population 2 ends Population 1’s epidemic.

Table 11 illustrates how isolation of the vaccinated and unvaccinated populations from each other interacts with the discontinuation of NPIs. The most remarkable part of this table is the rightmost column, which shows the results of completely abandoning NPIs. The vaccinated Population 1 suffers a case rate of 68 cases/thousand/year if it is interacting freely with Population 2, but a case rate of essentially zero when it is isolated from Population 2. On the other hand, Population 2’s case rate more than doubles when it is cut off from Population 1. The source of the Population 1 infections is Population 2. Also, it is apparent that the bulk of 1’s transition to such a disease-free state occurs between an interaction parameter of 0.1 and 0. In the rightmost column of Table 11, merely going from a 0.0 to a 0.1 interaction with Population 2 is sufficient to multiply Population 1’s case rate by a factor of approximately 6 billion. Even though Population 1 has abandoned NPIs, the vaccine allows it to control the disease that originates within itself. However, its case rate increases markedly when it is exposed even slightly to Population 2.

Also in Table 11, compare the Population 1 case rates in the Interaction = 1.0 row and in the Interaction = 0.0 row. In the top row, if Population 1 is interacting randomly with Population 2, Population 1’s case rate multiplies by about 70 times as the lockdown parameter changes from 0.5 to 0.0. But if interaction = 0.0, Population 1’s case rate remains very low and hardly changes as the lockdown parameter goes to zero. Population 1 can control its own disease even with no NPIs if it is independent, but not if it is exposed to Population 2 infections.

Since a zero interaction with the unvaccinated is not possible in the real world, the results above are also an endorsement of the maxim that “no one is safe until everyone is safe.” If Population 1 wants to keep itself disease-free, rather than trying to obtain better vaccines for itself, it would be far more productive to vaccinate Population 2 and convince it to maintain its NPIs.

The arrival of a more transmissible variant can drastically increase the post-vaccine case rates. It is remarkable how small the increase in R_0_ can be and still have a strong effect, even if vaccination with a high-efficacy vaccine is proceeding. In Table 12, even an increase of R_0_ from 2.3 to 3.0 increases the post-vaccine case rate by a factor of 5.8 for a fast vaccination rate of 0.5%/day, and by 6.1 times for a vaccination rate of 0.25%/day. If the increase is from an R_0_ of 2.3 to 4.0, the case rate goes up by 48.3 times for the 0.5%/day vaccination rate and by 36.0 times for the 0.25%/day rate. If we accept Fauci’s suggestion that case rates should be below 10 cases/thousand/year in the US, even an R_0_ of 4.0 would require a vaccination rate greater than 1%/day. US experience in 2021 tends to suggest that US vaccination rates will be between 0.2%/day and 0.5%/day. Table 12 predicts that with realistic vaccination speeds, the dominance of a variant with an R_0_ of 4.0 or higher will lead to post-vaccine case rates higher than anything seen in Figure 6, even if vaccination is going on as quickly as possible with an “ideal” vaccine.

When considering the vaccination of a population affected by a high-transmission variant, one might imagine that the disease and the vaccine are in a race to convert susceptibles to either new cases (by contracting the disease) or vaccine-induced immunes. The relative advantage of the vaccine falls in Table 12 as the R_0_ increases. With an R_0_ of 3.0, a vaccination rate of 0.5%/day decreases the case rate by a factor of almost 45 from the rate seen with no vaccine; with an R_0_ of 6.0, the same decrease is only by a factor of 1.7. The disease is spreads so quickly that the vaccine cannot keep up.

Table 13 makes a familiar point: the damage done by high-transmission variants can be reduced if the duration of immunity is longer. A variant with an R_0_ of 4.0 produces a post-vaccine case rate of 365.49 cases/thousand/year when immunity lasts six months. When immunity lasts 24 months, the case rate will be held to 18.24 cases/thousand/year. However, duration of immunity has its limits--if R_0_ = 6.0, even 24 months of immunity will allow a case rate of 152.92 cases/thousand/year.

Table 14 explores the advantages of vaccinating a larger percentage of the population. For a variant with an R_0_ of 4.0 or less, going from vaccinating 50% of the population to vaccinating 90% can cut the case rate by a factor of almost 20. However, the higher the new variant’s R_0_ is, the less advantageous this ratio gets. For an R_0_ of 6.0, going from 50% vaccinated to 90% vaccinated only reduces the case rate by a factor of 2.2, leaving it very high. Even 100% vaccination at the rate of 0.5% of the population/day probably cannot extinguish the epidemic, but it can hold the case rate of a high-transmission variant to a more moderate level.

Table 15 looks at the results of vaccinating a larger percentage from another point of view. For various percentages of Population 1 and Population 2, what vaccination speed (%/day) is necessary to hold the Population 1 case rate at the suggested level of 10 cases/thousand/year? Recall that a generous estimate of the mean vaccination rate in the US up to November of 2021 is 0.34%/day. Compare the bottom line of Table 15 with the top line. If the portion of the population that accepts vaccination is only 50%, the necessary vaccination rate is a fast but attainable 0.58% of the population/day for R_0_ = 2.3. However, for higher R_0_s, the necessary vaccination rates rapidly become impossible, even reaching 25% of the population *per day* for R_0_ = 6.0. On the other hand, the top line of the table shows that if 100% of the population can be vaccinated, much more modest vaccination rates can handle new variants with R_0_s at least up to 5.0.

The simulations suggest four possible solutions to the development of a high-transmission variant.

First, Table 13 shows that, as seen in other simulations in this report, a longer duration of immunity dramatically lowers case rates, so producing a vaccine that produces a longer duration of immunity would be a critical goal. However, Table 3 shows that even a vaccine that produces permanent immunity cannot eradicate an epidemic if vaccination speed is low and vaccine hesitancy is high.

The second approach to coping with a more transmissible variant, and the one being used in the US in 2021, is to vaccinate the largest possible fraction of the population. Table 15 shows that the larger the fraction that is vaccinated, the slower the speed of vaccination needs to be.

The third solution to high-transmission variants is to increase the vaccination speed. Vaccine hesitancy and slow vaccination rates appear to be the weakness of the US vaccination effort. The results in this report show that a fast vaccination rate allows the vaccine to compensate for a short duration of immunity, weakening of NPIs, vaccine hesitancy by a fraction of the population, and the spread of more transmissible variants. On the other hand, even the imaginary “ideal” vaccine simulated here can be rendered ineffective by a slow rate of vaccine distribution. The simulations predict that a slow rate of completed vaccination (such as the 0.25%/day rate that was seen in the US in summer of 2021) will allow high-R_0_ case rates to go higher than the worst values seen in the epidemic at the end of 2020.

The last line of defense against high-transmission variants is NPIs. NPIs such as masking and restrictions on contacts with others cannot be ignored in the post-vaccine era. Probably the most surprising of the results of this study was that even if the pathogen R_0_ remains at 2.3 and the vaccination rate remains at 0.5%/day, dropping the “lockdown” parameter from 0.5 to 0.25 will more than triple the case rate, and dropping the lockdown parameter to zero (totally abandoning NPIs) will increase the case rate by about 70 times (Table 6). This is due to cases that originate in unvaccinated Population 2. Table 16 contains even more disturbing data. If the pathogen R_0_ increases from 2.3 to values ranging from 3.0 to 6.0, dropping the lockdown parameter to zero from 0.5 will raise the case rates from single digits to hundreds of cases/thousand/year. On the other hand, those very high case rates can be lowered if the lockdown parameter can be increased from 0.5 to higher values (up to 0.8). Given popular resistance to lockdowns, a sustained increase in the lockdown parameter is probably an unrealistic hope, but the main message is that NPIs are not a relic of the pre-vaccine era. Especially when high-transmission variants are present, NPIs are a vital part of the battle against the pathogen. Vaccines cannot do the job alone unless the speed of vaccination is unrealistically fast. And Table 12 shows that even with the lockdown parameter maintained at 0.5, moderate vaccination rates cannot keep the post-vaccine case rate from reaching hundreds of cases per thousand per year when high-transmission variants are present.

Table 17 summarizes the model’s predictions of post-vaccine case rates for an “optimistic” and a “pessimistic” set of parameters. The optimistic case relies on 75% of the population being vaccinated, a fast vaccination rate, a small loss of interest in getting vaccine boosters in future years, only a small decline in NPIs after 500 days, and especially on a new variant with an R_0_ of only 3.0. The pessimistic alternative imagines that the percent of the population fully vaccinated is only slightly above 50%, there are severe declines in NPIs and seeking of booster vaccines, and that the new variant has an R_0_ of 5.0.

The results of these simulations are distinctly different. For one year of vaccine-induced immunity, the “optimistic” outcome produces an average combined case rate of 25.26 cases/thousand/year over the four post-vaccine years, with Population 1’s rate being 14.18. Population 1 is above the proposed limit of 10 cases/thousand/year, but only slightly. The optimistic scenario predicts that an average member of the population has a 2.5% chance of a COVID case each year of the post-vaccine era. On the other hand, the “pessimistic” scenario anticipates a case rate of 584.68 cases/thousand/year. This is twice as high as any national monthly case rate the US has experienced in the pandemic so far. It produces nearly a 60% chance of a case each year for an average member of the population. Even vaccinated Population 1 in the pessimistic scenario has a case rate of 460.47 cases/thousand/year because its 55% is sharing space with 45% unvaccinated Population 2, and also because Population 1’s interest in vaccination is dropping by 50% per year after the first year.

Figures 8 and 9 also show a sharp difference in the time course of the infection in the two scenarios, which are not disclosed by the case rates alone. Fig. 8 shows that after the initial peak, the “optimistic” infection rate settles down to a low, endemic level. In the “pessimistic” case in Fig. 9, the infection rate is low most of the time, but is punctuated by brief, explosive peaks. Remembering that the R_0_ in the pessimistic case is 5.0, repeated peaks can be explained by an expiration of immunity that leads to the release of a large number of susceptibles, a rapid disease spread through these susceptibles, and then an equally rapid decline of new cases as herd immunity is reached. Because of the large number of individuals getting disease-induced immunity at nearly the same time as the peak occurs, their immunity will expire simultaneously a year later, triggering another sharp peak. Once the susceptibles are generated by the expiration of immunity, it will be a race between the vaccine and the disease. Given the fact that the “pessimistic” vaccination rate is only 0.2%/day, a high-R_0_ pathogen will win the race and convert most of the susceptibles into new cases.

As before, duration of immunity has a strong effect. For a pessimistic scenario for Population 1, six months of immunity produces case rates about twice as high as one year, and two years of immunity produces case rates about 1/4 as large as one year. The optimistic scenario shows an even more powerful effect of the duration of immunity. Going from 6 to 12 months of immunity can cut case rates for Population 1 by a factor of 11, and by another factor of 4 as we go from 12 months of immunity to 24 months of immunity.

The major factor producing such high case rates is the higher R_0_ of the new variant. If the “optimistic” one-year simulations had kept their R_0_ at 2.3 instead of increasing it to 3.0, the post-vaccine case rate for Population 1 would have been 1.50 rather than 14.18. On the other hand, if the “optimistic” one-year simulations had kept all parameters the same as in Table 17 but had raised their pathogen’s R_0_ to 5.0, the Population 1 case rate would have been 186.23 instead of 14.18. Ironically, the “pessimistic” simulation is less sensitive to R_0_ because of its weak NPIs and slow vaccination rate. If the “pessimistic” simulation had kept all its parameters the same but had dropped its new variant R_0_ from 5.0 to 3.0, its Population 1 case rate would have been 306.43 instead of 460.47, very high in both cases.

Clearly, a highly-transmissible variant is a difficult public health challenge. The challenge cannot be met by an excellent vaccine alone--both “optimistic” and “pessimistic” populations in Table 17 had an excellent vaccine. The vaccine must be coupled with a rapid rate of vaccine distribution and strenuous use of NPIs. If the one-year “pessimistic” scenario in Table 17 had kept a final R_0_ of 5.0, but had raised its vaccination rate to 1% per day, allowed no decline in vaccination-seeking after the first year, and maintained a lockdown factor after 500 days of 0.75 instead of 0.2, the vaccine-era case rate for Population 1 would have been held to 27.05 cases/thousand/year, about 1/17 of the case rate in Table 17. When combating a high-R_0_ variant, success is still possible, but only with fast vaccination and stringent attention to NPIs.

We must remember that these are the results with an “ideal” vaccine that produces same-day sterilizing immunity against any variant. If the vaccine had realistic limitations, such as failure against a new variant, both the “optimistic” and “pessimistic” results would produce even higher case rates. So it might be said that the tables in this report are not a prediction of what will happen in reality; these case rates are the *best* that can be expected.

On the other hand, there is reason to accept these results with caution. First, we must consider a general weakness of simulations. Assuming that they include the proper relationships and parameters, simulations estimate what would happen if a set of conditions is continued for the length of the simulation. We certainly hope that if the pessimistic scenario were in progress and case rates were escalating drastically, the parameters of the system would react and change--NPIs would be restored, vaccination speeds would go up, and some Population 2 individuals would shift to Population 1. These responses would moderate the results.

A related objection is the assumption that the members of the whole population are interacting randomly with one another. The complex history of COVID-19 in the US has many examples where cases would flare up in a region, but then die out without spreading nationwide. The popular press has mentioned a “two-month COVID cycle,” in which cases tend to increase for two months and then recede, for unknown reasons (Leonhardt and Wu, 2021). This phenomenon may be caused by localized epidemics that increase until enough susceptibles have been immunized so that case rates fall again. Also, if cases are high in a region, the local population may resume NPIs so that the spread is reduced before it reaches its expected full extent.

Nevertheless, the failures of the ideal vaccine in this report can be instructive. Assuming that we want to hold a COVID epidemic to fewer than 10 cases/thousand/year in the post-vaccine era, these results predict that that even an “ideal” vaccine will find it hard to achieve that goal when:

a. the speed of vaccination is less than 0.1% of the population per day; or
b. the percent of the population willing to accept the vaccine is less than 50%; or
c. the duration of vaccine-induced immunity is shorter than 9 months; or
d. fading of interest in getting the vaccine is 50% per year or more; or
e. NPIs weaken or are completely abandoned after the advent of the vaccine; or
f. a new variant developing after the initial outbreak has an R_0_ of 3.0 or 4.0, except when more than 90% of the population accepts the vaccine; or
g. the new variant has an R_0_ of 5.0 or greater.

When multiple adverse conditions on the list above are true, even an “ideal” vaccine may be unable to keep the case rate below 10 cases/thousand/year.

## Data Availability

All data produced in the present work are contained in the manuscript.

